# Generating clinical-grade pathology reports from gigapixel whole slide images with HistoGPT

**DOI:** 10.1101/2024.03.15.24304211

**Authors:** Manuel Tran, Paul Schmidle, Sophia J. Wagner, Valentin Koch, Brenna Novotny, Valerio Lupperger, Annette Feuchtinger, Alexander Böhner, Robert Kaczmarczyk, Tilo Biedermann, Nneka I. Comfere, Ruifeng (Ray) Guo, Chen Wang, Kilian Eyerich, Stephan A. Braun, Tingying Peng, Carsten Marr

**Affiliations:** Helmholtz AI, Helmholtz Munich, Neuherberg, Germany; School of Computation, Information and Technology, Technical University of Munich, Munich, Germany; Dermatology Department, University Hospital Münster, Münster, Germany; Institute of AI for Health, Helmholtz Munich, Neuherberg, Germany; Department of Quantitative Health Sciences, Mayo Clinic, Rochester, Minnesota, United States; MLL Munich Leukemia Laboratory, Munich, Germany; Core Facility Pathology and Tissue Analytics, Helmholtz Munich, Neuherberg, Germany; Department of Dermatology and Allergy, Technical University of Munich, Munich, Germany; Dept. of Dermatology and Laboratory Medicine & Pathology, Mayo Clinic, Rochester, Minnesota, United States; Department of Laboratory Medicine and Pathology, Mayo Clinic, Jacksonville, Florida, United States; Department of Dermatology, Medical Center, University of Freiburg, Freiburg, Germany; Department of Dermatology, Medical Faculty, Heinrich-Heine University, Düsseldorf, Germany

## Abstract

Histopathology is considered the reference standard for diagnosing the presence and nature of many malignancies, including cancer. However, analyzing tissue samples and writing pathology reports is time-consuming, labor-intensive, and non-standardized. To address this problem, we present HistoGPT, the first vision language model that simultaneously generates reports from multiple pathology images. It was trained on more than 15,000 whole slide images from over 6,000 dermatology patients with corresponding pathology reports. The generated reports match the quality of human-written reports, as confirmed by a variety of natural language processing metrics and domain expert evaluations. We show that HistoGPT generalizes to six geographically diverse cohorts and can predict tumor subtypes and tumor thickness in a zero-shot fashion. Our model demonstrates the potential of an AI assistant that supports pathologists in evaluating, reporting, and understanding routine dermatopathology cases.

---

Histopathology stands as the clinical gold standard for the diagnosis of a wide range of diseases, including malignant cancers and inflammatory disorders^1^. It involves the examination and interpretation of diseased tissues and cells under a microscope^2^ according to widely accepted international guidelines that ensure accuracy, consistency, and objectivity. The analysis results are then summarized in a comprehensive pathology report that serves as the basis for the communications with clinicians for further testing and treatment. Creating such reports, however, is time-consuming, labor-intensive, and non-standardized^3^. For some tumors, such as basal cell carcinoma, an experienced pathologist can make a diagnosis in seconds, but it takes considerably longer to dictate or type the microscopic findings. With the number of cancer cases increasing and the number of pathologists decreasing in many countries^4^, patient turnaround times are likely to worsen in the future.

Artificial intelligence (AI) has the potential to ease the burden on pathologists in their daily workflow by handling common and uncomplicated cases. Advanced systems like deep neural networks^5^ excel at vision tasks such as cancer classification^6^, tissue segmentation ^7^, survival prediction^8^, and biomarker detection^9^, and are typically applied to digitized microscope slides, also known as whole slide images (WSIs). Rather than replacing pathologists, AI is generally viewed as a tool and complement to other medical tests^10^.

Currently, there are two approaches to computational pathology. Patch-level approaches use a single crop of a WSI (called an image patch), ranging from 224 x 224 pixels to 1024 x 1024 pixels, to generate an output^11^. Thus, by design, these patch-level approaches ignore up to 99% of the entire tissue, miss potentially diagnostically relevant areas, and cannot be applied to tasks that require the full context of the entire tissue sample (e.g., tumor thickness prediction). Slide-level approaches, on the other hand, aggregate the information from patch-level approaches into a slide-level representation that can be used in downstream tasks like biomarker prediction^9^. A recent research direction is to extend the capabilities of such methods by including text data. Contrastive vision language models in pathology, such as PLIP^16^ and CONCH^19^, align text and images at the patch level. They are zero-shot learners, which means they can solve downstream tasks they have never seen before like subtyping cancers. However, they cannot generate detailed text to describe the input image. Generative vision language models such as Med-PaLM M^14^, LLaVA-Med^17^, or PathChat^18^ can generate text, but only at the patch level for small image regions up to 1024 x 1024 pixels. This means that none of the existing medical foundation models can generate reports from an entire pathology image at full resolution, let alone from multiple pathology images.

Here, we present HistoGPT, the first vision language model to generate histopathology reports from gigapixel WSIs (see Fig. 1). Given multiple tissue sections from the same patient at up to 20x magnification, HistoGPT uses a vision foundation model (VFM) to extract meaningful features from the images and combines them with a large language model (LLM) via cross-attention mechanisms to generate the final report. The generated report describes the tissue composition, cellular subtypes, and potential diagnosis. In addition, users can interact with the model through various prompts (“Expert Guidance”) to extract additional information such as tumor subtypes and tumor thickness predictions (see Fig. 1B). The output text is fully interpretable because HistoGPT provides saliency maps that highlight the corresponding image regions for each word or phrase in the generated text (see Fig. 1C).

**Figure 1:**
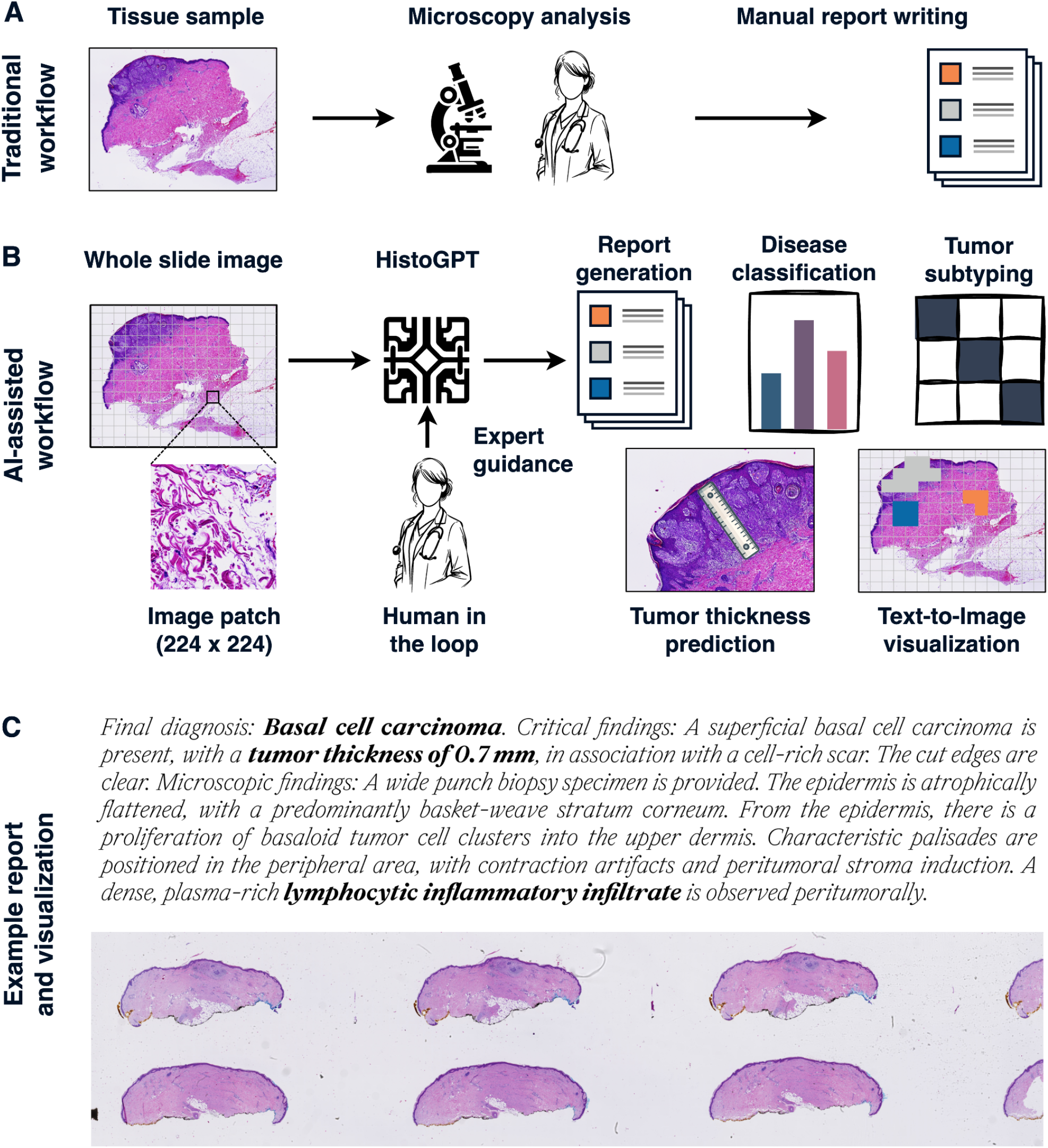
HistoGPT, a vision language foundation model for dermatopathology. (A) Traditionally, pathologists analyze tissue samples from patients under a microscope and summarize their findings in a comprehensive pathology report. This manual process is time-consuming, labor-intensive, and non-standardized. (B) In our proposed AI-powered clinical workflow, pathologists work alongside HistoGPT. It generates human-level written reports, provides disease classification, discriminates between tumor subtypes, predicts tumor thickness, and returns text-to-image gradient-attention maps that provide model explainability. All this serves as a second opinion for the pathologist, who can use the output of HistoGPT as a general overview and first draft for the final report. (C) We provide an example of such a generated report for a basal cell carcinoma case from our external validation cohort.

To train HistoGPT, we collect a large multimodal skin histology dataset from the Department of Dermatology at the Technical University of Munich with 15,129 paired WSIs and pathology reports from 6,705 patients written by board-certified pathologists. To validate HistoGPT, we are using one internal and six external test cohorts covering different scanners, medical procedures, and countries. To our knowledge, we also provide the first evaluation of a pathology vision language model by an international team of board-certified pathologists from a clinical perspective. In this regard, the reports generated by HistoGPT are often found to be highly consistent with both the reference report and histologic specimens for the most common diseases. We are releasing HistoGPT in different sizes as an end-to-end deep learning pipeline that can be deployed on local machines. As a result, users can select and fine-tune a copy of our machine learning algorithm according to their needs.

## Results

### HistoGPT is the first slide-level model that learns from image and text

HistoGPT is a family of models with three configurations (small, medium, and large), each consisting of two components (see Fig. 2A): a vision module and a language module. The vision module is based on the patch encoder CTransPath (CTP)^22^ for the small and medium models and UNI^23^ for the large model. The former is a lightweight Swin Transformer^24^ trained on over 32,000 WSIs from TCGA^25^ and PAIP^26^ using a semantically guided contrastive learning algorithm^27^. The latter is a much larger Vision Transformer^28^ trained on over 100,000 WSIs from 22 major tissue types using self-distillation and masked modeling^29^. Our language module reuses BioGPT^20^, an autoregressive generative model based on the Transformer^30^ decoder architecture of GPT-3^31^ trained on 15 million biomedical articles from PubMed.

**Figure 2:**
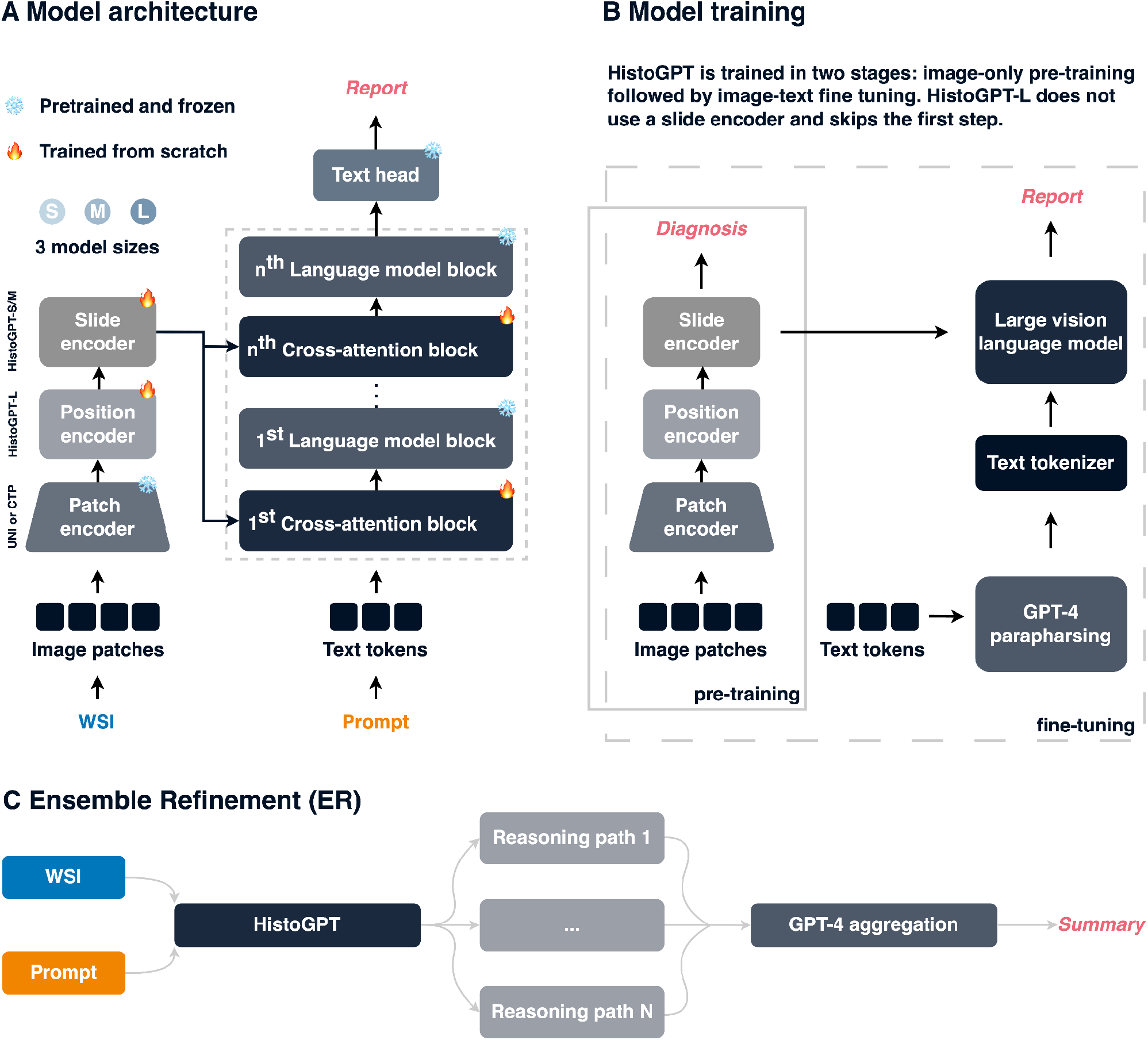
HistoGPT learns simultaneously from vision and language to generate clinical-grade histology reports from whole slide images. (A) HistoGPT is available in three sizes (*S*mall, *M*edium, and *L*arge). It consists of a patch encoder (CTP for HistoGPT-S and HistoGPT-L as well as UNI for HistoGPT-L), a position encoder (used only in HistoGPT-L), a slide encoder (used only in HistoGPT-S and HistoGPT-M), a language model (BioGPT base for HistoGPT-S as well as BioGPT large for HistoGPT-M and HistoGPT-L), and gated cross-attention blocks. Specifically, HistoGPT takes a series of whole slide images (WSIs) as input and outputs a written report. Optionally, users can query the model for additional details using prompts such as “The tumor thickness is”, and the model will complete the sentence, e.g., “The tumor thickness is 1.2 mm”. (B) We train HistoGPT in two phases. In the first phase, we pre-train the vision module of HistoGPT using multiple instance learning (MIL). In the second phase, we freeze the pre-trained layers and fine-tune the language module on the image-text pairs. Since HistoGPT-L does not use a slide encoder, it skips the first step. To prevent the model from overfitting on the same sentences, we apply text augmentation. This is done using GPT-4, a general-purpose large language model that faithfully paraphrases the medical notes. (C) During deployment, we can optionally use an advanced inference method called Ensemble Refinement (ER). Here, the model stochastically generates multiple possible pathology reports using temperature sampling to capture different aspects of the input image. An aggregation module (GPT-4) then combines the results to provide a more complete description of the underlying case.

For the small and medium models, we sample image features from the vision module with a custom pre-trained (see Fig. 2B) slide encoder based on the Perceiver Resampler^32^ and integrate its outputs into the LLM via interleaved gated cross-attention (XATTN) blocks^33^. Only these new XATTN blocks are trained from scratch. In this way, we endow HistoGPT with existing visual and linguistic domain knowledge, which is crucial for tackling the challenging problem of generating histopathology reports from entire WSIs. Similar to Flamingo^33^, we freeze the parameters of all pre-trained modules during optimization to further reduce the computational cost and to avoid catastrophic forgetting of the inductive biases encoded in the learned weights.

The large model skips the Perceiver Resampler. This is computationally more expensive because we work directly with all high-dimensional feature vectors, but we do not lose any information due to compression. The large model uses a graph convolutional network^34^ to encode slide-level position information, which we remove for the small and medium models due to its space and time complexity. Due to the computational resources required to run the large model, we report most of our experiments for the small and medium models, which are much faster and more likely to be used in practice.

A language model predicts a probability distribution over a vocabulary. The next word in a text is randomly selected based on a combination of top-p and top-k sampling. Once the first few words have been chosen, the outline of the report is roughly pre-determined. To avoid being locked into a fixed text structure, we use an advanced inference method called Ensemble Refinement, introduced in Med-PaLM 2^35^, to randomly sample multiple reports – each focusing on slightly different aspects of the WSI (see Fig. 2C). This sampling allows us to thoroughly search the model distribution and generate a wide variety of medical reports, maximizing the likelihood of including all important observations. A general-purpose LLM such as GPT-4^21^ is then used to aggregate all the bootstrapped reports.

### HistoGPT generates human-level pathology reports for common diseases

We train HistoGPT on 15,129 whole slide images from 6,705 dermatology patients with corresponding pathology reports from a real-world medical cohort (see Fig. 3A). This internal dataset contains 167 skin diseases of varying frequency and has a total size of 10 terabytes. To assess the impact of model architecture and parameter size, we train and evaluate four versions of HistoGPT: HistoGPT-S, HistoGPT-M, HistoGPT-L, and HistoGPT-L-ER as described above. In the following experiments, we use HistoGPT in “Expert Guidance” mode, where the model is prompted with the correct diagnosis, simulating a pathologist who is confident in the WSI assessment but wants to leave the work of writing a draft to an AI assistant (see Fig. 3B).

**Figure 3.**
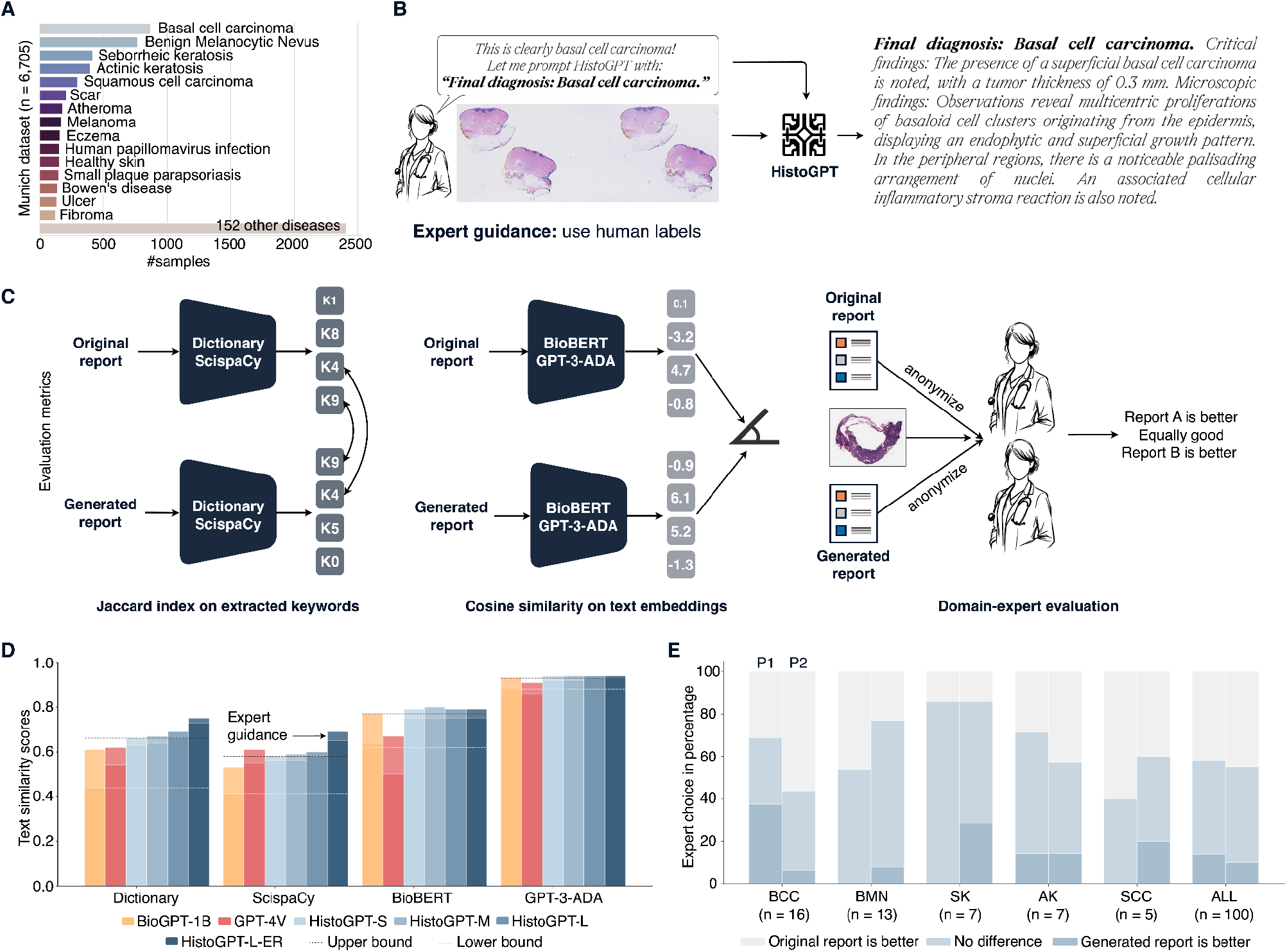
HistoGPT generates human-level pathology reports of skin diseases. (A) Our internal Munich dataset is a real-world cohort of 6,705 patients with 167 skin diseases from the Department of Dermatology at the Technical University of Munich. It includes malignant cases such as basal cell carcinoma (BCC, n = 870) and squamous cell carcinoma (SCC, n = 297); precursor lesions such as actinic keratosis (AK, n = 396) as well as benign cases such as benign melanocytic nevus (BMN, n = 770) and seborrheic keratosis (SK, n = 412). We divide the patient-level dataset into a training set and a test set using a stratified 85/15 split. (B) Through years of experience, pathologists are often able to make a diagnosis at first glance. Instead of writing a pathology report themselves, they can use HistoGPT in “Expert Guidance” mode by giving the model the correct diagnosis to complete the report. (C) We evaluate the model’s performance using four semantic-based machine learning metrics: (i) match critical medical terms extracted from the original text with the generated text using a dermatology dictionary; (ii) use the same technique but with ScispaCy, a scientific name entity recognition tool, as the keyword extractor; (iii) compare the semantic meaning of the original and generated reports by measuring the cosine similarity of their text embeddings generated by the biomedical language model BioBERT; (iv) use the same technique but with the general purpose large language model GPT-3-ADA for text embedding. (D) In “Expert Guidance” mode (translucent colors), HistoGPT-L-ER (HistoGPT-L with Ensemble Refinement) surpasses BioGPT-1B and GPT-4V on the two text accuracy metrics Dictionary and ScispaCY and is also better on the two text similarity metrics, BioBERT and GPT-3-ADA (see Methods for details). (D) Two independent external board-certified pathologists (P1 left and P2 right) evaluated 100 original and generated reports together with the corresponding WSI in a randomized, blinded study. For BCC, P1 found that 38% of the generated reports described the WSI better than the original report. In 31% of cases, both reports performed equally well, while in 31% of cases, the original report was preferred. In 58% (P1) and 55% (P2) of all cases, the pathologists did not prefer the original report to the generated one.

Currently, no deep learning model can generate a histopathology report from an entire WSI, let alone a series of WSIs for patients with multiple tissue sections cut from the tissue block. Therefore, we compare the generated reports with those of text-only and patch-only architectures. For the former, we choose the domain-specific language model BioGPT-1B, fine-tuned on our internal cohort. For the latter, we rely on the multimodal foundation model GPT-4V(ision)^21^, which takes low-resolution images of size 2000 x 768 as input. We introduce two additional baselines to further validate that HistoGPT does not simply memorize and repeat sentences from the training set: A lower baseline, where we select two random reports with arbitrary diagnoses; and an upper baseline, where we compare two random reports with the same diagnosis (see Methods for more details).

We evaluate the performance of the models using four semantic-based machine learning metrics as well as two blinded domain expert evaluations (see Fig. 3C). In “Expert Guidance” mode, HistoGPT-L captures on average 69% of all dermatological keywords^56^ from the original pathology reports (see Fig. 3D), outperforming alternative language models such as BioGPT-1B and GPT-4V by at least 7%. This gap is reduced to 5% with the lightweight HistoGPT-S and HistoGPT-M. HistoGPT-L-ER further improves the Jaccard index to 75%, which is 9% above the upper baseline. A similar trend is observed when ScispaCy^36^ is used as a keyword extractor (see Fig. 3D). All variants of HistoGPT produce text with high cosine similarity to the ground truth, as indicated by the embeddings provided by BioBERT^37^ and GPT-3-ADA^31^ (see Fig. 3D).

Overall, “Expert Guidance” is the recommended modus operandi for HistoGPT because it allows a pathologist to work interactively with the model while improving the quality of the report compared to the unguided mode (see Fig. 3D, translucent vs opaque columns). We also evaluate all models using traditional syntax-based measures (BLEU-4, ROUGE-L, METEOR, and BERTscore). The relatively low syntax-based scores (see Supplementary Table: Automatic Report Scoring) combined with the high semantic-based scores (see Fig. 3D) further confirm our hypothesis that HistoGPT is not overfitting the training set by simply repeating common phrases and medical terms, but is deeply rooted in the input image.

To evaluate the content of the generated reports from an expert perspective, we conducted a blinded study in which we randomly selected 100 cases from our Munich test split, generated a report for each patient in “Expert Guidance” mode, and paired it with the original human-written report. The two reports were then randomly shuffled and anonymized. Two independent board-certified pathologists, who were neither involved in the construction nor annotation of the Munich cohort, were given the original WSIs and asked to identify the report that best described each case, with the option of selecting “no difference” if both were deemed equally accurate. Ensemble Refinement was not used in this study to avoid easy identification of the GPT-4 generated summary. For the five largest diagnostic classes (basal cell carcinoma (BCC), benign melanocytic nevus (BMN), seborrheic keratosis (SK), actinic keratosis (AK), squamous cell carcinoma (SCC), see Fig. 3A), we found moderate agreement between the two pathologists. Analyzing the results for each class separately, we found that Pathologist 1 overwhelmingly preferred the AI or found the AI and human report similarly good in about 70% of the BCC cases. Pathologist 2, on the other hand, preferred the AI-generated report for BMN 80% of the time. The AI-generated report for SK is preferred by both pathologists 90% of the time. Across all 100 report pairs, both pathologists found no difference between the generated and human reports about 45% of the time and preferred the AI-generated reports about 15% of the time (see Fig. 3D).

According to a post-analysis provided by the two pathologists, after about 20 cases, they were able to tell which report was likely generated by the AI. The AI-generated text tends to be more structured and comprehensive. It includes more observations that are informative but not always necessary for the final diagnosis (for example, the AI mentioned a bystander cyst that was irrelevant to the diagnosis of BCC). Overall, there are some interesting cases worth mentioning: In one interesting case, there was a disagreement between the ground truth diagnosis and Pathologist 1 – resulting in both AI and human reports being disputed. In another case, a slide was incorrectly annotated by the human, but the AI still provided the correct report. In one case, the AI failed to detect small or unusual objects, such as a scabies mite. In one slide, the model mistook erythrocytes for eosinophils. However, these two cell types were difficult to distinguish due to the quality of the slide. Pathologist 1 mentioned that about 10 human reports were preferred because the reported tumor thickness seemed more accurate than in the generated report, but the text itself was equally good. After adjusting for this, and including only reports where “Expert Guidance” and model prediction agreed, the pathologist preferred the AI report or was indifferent 80% of the time (see Supplementary Figure 1). In retrospect, the model was described as having the skill level of a novice pathologist. Notably, this was achieved with only 5K training samples, which is small by LLM standards.

### HistoGPT accurately predicts diseases in geographically diverse cohorts

To demonstrate from another perspective that HistoGPT has effectively learned to encode medical knowledge, we extract the predicted diagnosis from the generated reports, calculate the classification accuracy, and compare the results with state-of-the-art multiple instance learning (MIL) approaches. For this purpose, we run HistoGPT without “Expert Guidance”, i.e. we only prompt the model with the phrase “Final diagnosis” instead of “Final diagnosis: [expert label]” and let it make a diagnostic decision on its own (see Fig. 4A).

**Figure 4.**
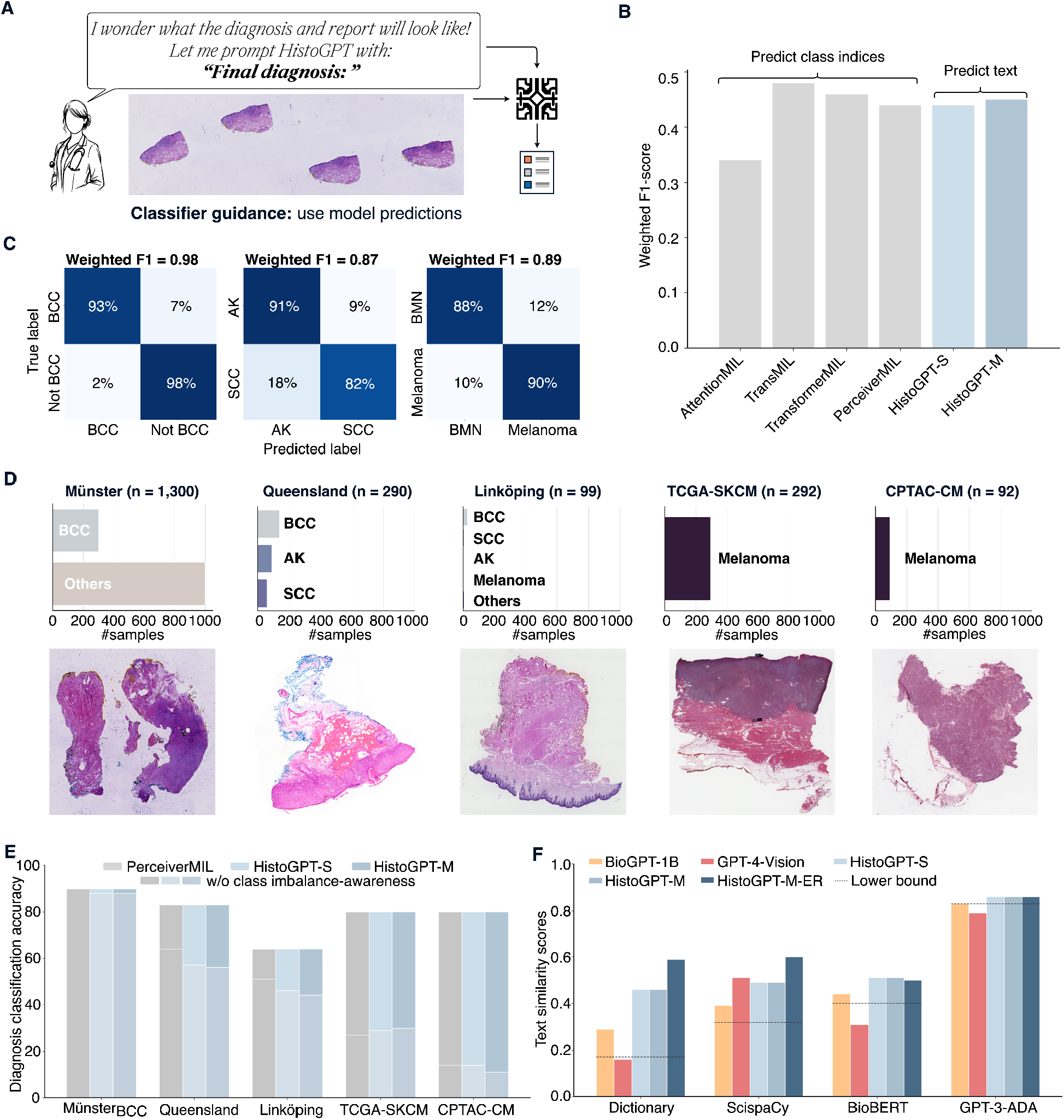
HistoGPT accurately predicts diseases in-domain and out-of-domain without human guidance. (A) In the absence of a human-in-the-loop, HistoGPT predicts the patient’s diagnosis on its own and generates the corresponding pathology report. (B) On the internal Munich test set, HistoGPT is comparable to state-of-the-art classification models in predicting over 100 dermatological diseases, even though the model’s output is pure text. (C) HistoGPT (medium) discriminates malignant from benign conditions with high accuracy on the Munich dataset: basal cell carcinoma (BCC, n = 107) vs. other conditions (n = 621) with an accuracy of 0.98 and a weighted F1 score of 0.98; actinic keratosis (AK, n = 47) vs. squamous cell carcinoma (SCC, n = 33) with an accuracy of 0.88 and a weighted F1 score of 0.87; benign melanocytic nevus (BMN, n = 86) vs. melanoma (n = 21) with an accuracy of 0.89 and a weighted F1 score of 0.89. (D) We evaluate HistoGPT in five independent external cohorts covering different countries, scanner types, staining techniques, and biopsy methods. (E) Both PerceiverMIL and HistoGPT perform well on external datasets by conditioning them on the class distributions. (F) HistoGPT is able to produce highly accurate pathology reports, as indicated by the high keyword and cosine-based similarity scores on Münster. As in Figure 3C, the lower baseline compares two randomly selected reports.

Multiple instance learning methods such as AttentionMIL^12^, TransMIL^13^, and TransfomerMIL^9^ achieve relatively low weighted F1 scores (a measure that balances precision and recall across different classes, giving more importance to larger classes) between 0.34 and 0.48 on the Munich test set (see Fig. 4B). A major challenge for all these methods is that the training dataset is highly unbalanced, ranging from a handful of samples in the minority classes to several hundred samples in the majority classes (see Fig. 3A). Compared to PerceiverMIL, which achieves a weighted F1 score of 44% on the internal test set (see Fig. 4B), our much larger HistoGPT does not overfit and exceeds the performance of its vision module by 1%. Compared to the highly specialized AttentionMIL, TransMIL, and TransfomerMIL models, HistoGPT is competitive in terms of classification performance. It is important to note that, unlike MIL approaches, the output of HistoGPT is pure text and not integer class indices, highlighting the flexibility of a vision language model.

A challenging clinical question with a high therapeutic impact in dermatology is the differentiation of cancer from non-cancer. In routine diagnosis it is important to distinguish, for example, basal cell carcinoma (BCC) from other conditions; squamous cell carcinoma (SCC) from precancerous actinic keratosis (AK); and melanoma from benign melanocytic nevus (BMN). Unlike the previous classification task with over 150 classes, we now face a classification problem with only two alternatives. In this case, HistoGPT automatically calls a lightweight binary classifier to solve the task at hand (called “Classifier Guidance”, see Methods), overcoming the class imbalance problem from before. With HistoGPT, we achieve classification performance for the three clinical tasks with weighted F1 scores of 98%, 87%, and 89%, respectively (see Fig. 4C).

HistoGPT in “Classifier Guidance” mode also generalizes to previously unseen datasets and problems. We demonstrate this by evaluating HistoGPT on five external, publicly available cohorts from different countries, scanner types, staining protocols, and medical procedures such as shave biopsies, punch biopsies, and excisional biopsies (see Fig. 4D). While some of the cohorts include a variety of dermatologic diseases (Queensland and Linköping), some other cohorts (TCGA and CPTAC) include only melanoma cases, but can still be used to assess the accuracy of HistoGPT. Since F1 scores cannot be calculated for single-class datasets, we only present accuracy (see Fig. 4E) for all datasets and refer the reader to the Supplementary Tables for additional metrics. We retrain PerceiverMIL as a state-of-the-art classifier on the entire Munich cohort and compare its classification performance on the external datasets.

In the BCC subset of Münster, both PerceiverMIL and HistoGPT correctly identify BCC in 88% of cases (see Fig. 4E). In multi-class settings (Queensland with 3 classes and Linköping with 14 classes), we achieve accuracies of 83% and 64% and weighted F1 scores of 83% and 66%, respectively. The models also reliably discriminate melanoma from other types with accuracies of 80% and 90% in TCGA and CPTAC, respectively. For comparison, we also report the results of HistoGPT without class imbalance awareness (see Fig. 4E, light color bars). “Classifier Guidance” significantly improves the effectiveness and generalizability of the model across different external cohorts.

Of the five cohorts, only Münster (excluding the BCC subset) includes unstructured pathology reports. In contrast to the Munich reports, these reports contain only the critical findings and the final assessment (e.g., “Lichen planus-like keratosis (regressive solar lentigo/flat seborrheic keratosis), no evidence of basal cell carcinoma in the present biopsy.”) and thus lack the detailed microscopic description of the Munich training set. Since the critical findings include different classes not seen in Munich and are not available separately from the written text, it was not possible to extract individual class labels for classification. Nevertheless, we can calculate how much diagnostic information HistoGPT encodes by comparing the extracted keywords and measuring the cosine similarity (see Fig. 4F). HistoGPT captures nearly 60% of all biomedical keywords using our dermatology dictionary and the ScispaCy model, even though the ground truth was written in a completely different style and structure. HistoGPT also achieves high cosine similarity under BioBERT and GPT-3-ADA. Compared to a random report generated by BioGPT-1B and a grounded report given by GPT-4V, the text quality of these models is much lower compared to HistoGPT, with or without Ensemble Refinement (see Fig. 4F).

### HistoGPT predicts tumor thickness and tumor subtypes zero-shot

In the diagnosis of skin tumors, information about tumor thickness and assignment to a specific tumor subtype in the final report is critical. These parameters are well established in the literature: In basal cell carcinoma, tumor thickness is measured from the stratum granulosum of the epidermis to the deepest point of the tumor in millimeters, similar to the determination of the Breslow index in melanoma, while tumor subtype classification is based on the WHO guidelines^38^.

HistoGPT can predict both tumor thickness and tumor subtypes out-of-the-box and does not require additional reconfiguration or specification of tumor-specific parameters at any stage of training. By entering the prompt “The tumor thickness is” into HistoGPT, it will produce a prediction of the depth of tumor invasion without any fine-tuning. For the 94 samples in the internal Munich test set where tumor thickness was included in the original report, we measure a root mean square error (RMSE) of 1.8 mm and a significant Pearson correlation coefficient ρ of 0.52 (p = 9.7·10^−8^, two-sided test) for the predicted tumor thickness given by HistoGPT-M (see Fig. 5A). Binning the values to an interval with step sizes of 2 mm, 1 mm, and 0.5 mm yields accuracies of 64%, 38%, and 21%, respectively. In comparison, the predictions of the slide-level contrastive baselines (see Methods), HistoCLIP (RMSE = 4.35 mm, ρ = 0.006, p = 0.96) and HistoSigLIP (RMSE = 3.84 mm, ρ = 0.38, p = 0.002), correlate poorly with the ground truth and are far from HistoGPT in terms of quality (see Supplementary Figure 2). The patch-based contrastive baseline PLIP^16^, which is the state of the art in computational pathology, is even worse (RMSE = 2.78 mm, ρ = −0.18, p = 0.08), highlighting the importance of a slide-level approach. Accurate position encoding of each image patch seems to play an important role, as HistoGPT-L reduces the RMSE to 1.6 mm and increases the correlation to 0.67 (p = 9.1·10^−24^, see Supplementary Figure 4). Although only a fraction (n = 644) of the training reports contain a tumor thickness measurement, HistoGPT seems to have learned how to measure it accurately. This emergent behavior is known in the literature as zero-shot learning^39^.

**Figure 5.**
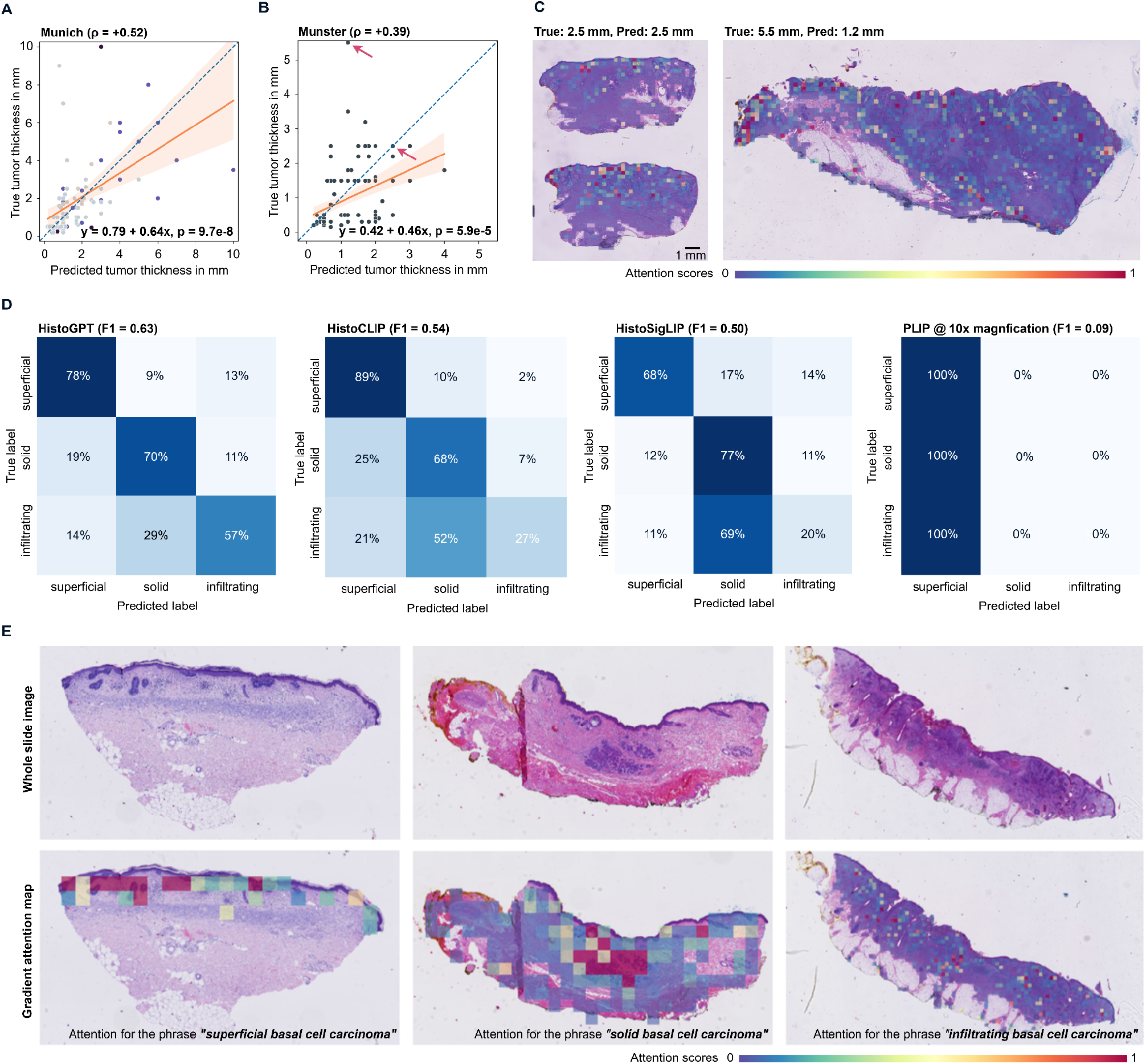
HistoGPT predicts tumor thickness and tumor subtypes in a zero-shot fashion and provides text-to-image visualization. (A) HistoGPT achieves high zero-shot performance in predicting tumor thickness on the internal Munich test set. The scatter plot is color-coded according to the classes in Figure 3A. (B) HistoGPT’s prediction is also highly correlated with the ground truth on the external Münster test set, even though it was obtained using a different measurement protocol. (C) Since HistoGPT is an interpretable AI system, we can fully understand its results. Here we show the two examples marked with a red arrow in Figure 5B. (D) On the basal cell carcinoma subset of the external validation set Münster, HistoGPT is the only slide-level model that correctly predicts infiltrative BCC in most cases. The patch-level model PLIP fails in this task, predicting all samples as superficial. (E) Given whole slide images of superficial, solid, and infiltrating BCC, HistoGPT correctly identifies their morphological structures as shown in the high attention regions for the respective text strings.

We analyze whether the zero-shot capability generalizes to other cohorts by looking at the never-seen BCC subset of the external Münster test set (see Fig. 5B), which was not used for training. For the samples with a ground truth tumor thickness measurement, we find a root mean square error of 0.98 mm and a significant Pearson correlation coefficient of 0.39 (p = 5.8·10^−5^). HistoGPT outperforms HistoCLIP (RMSE = 3.91 mm, ρ = −0.16, p = 0.1), HistoSigLIP (RMSE = 1.46 mm, ρ = 0.10, p = 0.3), and PLIP (RMSE = 1.43 mm, ρ = −0.04, p = 0.7), on this out-of-domain task (see Supplementary Figure 2).

Using gradient attention maps, we can gain insight into the reasoning steps behind each output. When estimating tumor thickness, HistoGPT correctly focuses on the tumor region (see Fig. 4C, left). However, it sometimes struggles to find the correct reference point (e.g., when the epidermis is torn or especially when it is ulcerated, see Supplementary Figure 3) or spatial orientation to start the measurements, even though it recognizes the tumor mass itself (see Fig. 4C, right).

We continue to explore the broader scope of zero-shot learning. Basal cell carcinoma is the most common type of malignant skin cancer. Although it is the majority class in the training set, the training set does not contain BCC subtypes as critical diagnoses. Therefore, BCC subtypes could not be used as labels during supervised pre-training. This information is only implicitly available as free text hidden in the report. Interestingly, HistoGPT is still able to extract the hidden information from the internal training set from Munich and apply the acquired knowledge in the external test set from Münster to discriminate between three major BCC subtypes (“superficial”, “solid/nodular”, and “infiltrating”) with a weighted F1 score of 63%, quantified by extracting the associated wordings from the generated reports (see Fig. 5D). As shown in the gradient attention maps (see Fig. 5E), HistoGPT correctly attends to the relevant architectural patterns within the histology slides that are the hallmarks of each BCC subtype.

This zero-shot capability highlights the adaptability of HistoGPT as a generative AI model, especially when compared to more traditional classifiers such as TransMIL, which are limited to predefined classes and thus cannot predict subtypes without re-training. We also compare its zero-shot performance to more advanced slide-level models such as HistoCLIP and HistoSigLIP. As contrastive methods, they overcome the inflexible structure of multiple instance learning approaches. Both achieve weighted F1 scores of 54% and 50%, respectively, but perform worse than HistoGPT, particularly in identifying infiltrating BCC (see Fig. 5D). Notably, infiltrating BCC is extremely important to identify in routine diagnostics, as this subtype tends to have a biologically much more aggressive growth pattern and a higher relapse rate. The patch-based vision language foundation model for pathology image analysis PLIP does not provide useful predictions for this zero-shot classification task. Surprisingly, PLIP is constant over the test set and predicts all specimens as either superficial or solid depending on the resolution. That is, at 5x and 10x magnification, PLIP predicts all cases as superficial; at 20x and 40x magnification, it predicts all images as infiltrating (see Supplementary Figure 5).

### HistoGPT generalizes to an independent real-world clinical evaluation

How does HistoGPT perform in a real-world dermatopathology setting? To answer this question, a subset of 52 cases consisting of 84 specimens was randomly selected from a one-week period at a tertiary medical center dermatology clinic (Mayo Clinic, USA, see Fig. 6). Cases were previously diagnosed by board-certified dermatopathologists with more than ten years of independent practice in academic centers with a digital pathology environment. Slides were scanned using a standard whole slide image scanner, and WSIs were viewed on a digital pathology image viewing platform in widespread use at the contributing authors’ institution. The 52 selected cases included 50 neoplastic epithelial lesions (including basal cell carcinoma, squamous cell carcinoma, actinic keratosis, verrucous keratosis, seborrheic keratosis, inverted follicular keratosis), four cases of nevus, four cases of dermatitis, two cysts, eight re-excisions (cases with “no residual” findings), two melanomas, one case of drug reaction (with generalized pustulosis), and 13 miscellaneous/other cases, for a total of 84 specimens (see Fig. 6B).

**Figure 6.**
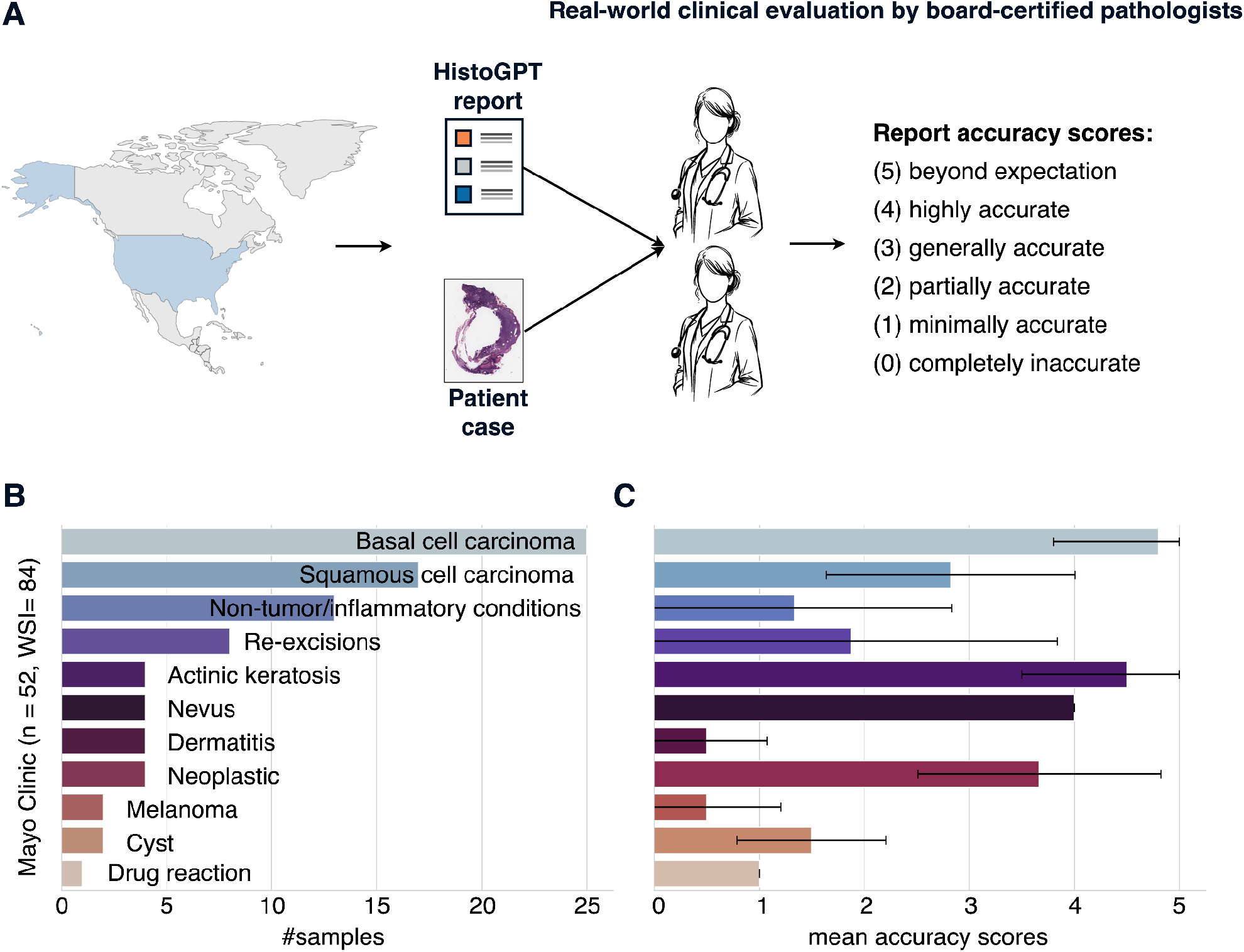
HistoGPT produces highly consistent and accurate pathology reports, as confirmed in a real-world clinical evaluation. (A) Skin biopsies were collected over the course of one week at the Mayo Clinic in the United States. A random subset (n = 52) was selected, and 84 digitized pathology images were processed by HistoGPT and scored by two independent board-certified pathologists in consultation with each other. Consistency and accuracy with respect to the underlying case were scored as follows: (5) beyond expectation, (4) highly accurate, (3) generally accurate with minor variations without clinical impact, (2) partially accurate with variations that could have a clinical impact, (1) minimally accurate, (0) completely inaccurate. In general, a score of 3 or higher indicates a diagnosis that is considered correct or within an acceptable range of subjectivity. (B) The class distribution represents cases from a one-week clinical period. (C) HistoGPT produces consistent and accurate reports for neoplastic epithelial lesions (mean score of 3.9) and struggles with classes with little training data.

The pathology reports generated by HistoGPT were unguided, i.e. neither “Expert Guidance” nor “Classifier Guidance” was used. They were analyzed by two independent board-certified dermatopathologists, ignoring differences in report format due to German vs. American standards. The quality of reports were scored as follows: (5) beyond expectation, (4) highly accurate, (3) generally accurate with minor variations without clinical impact, (2) partially accurate with variations that could have a clinical impact, (1) minimally accurate, (0) completely inaccurate (see Fig. 6A). A score of 3 or higher indicates a diagnosis that is considered correct or within an acceptable range of subjectivity.

According to the expert evaluation (see Fig. 6C), HistoGPT performed particularly well in diagnosing basal cell carcinoma (achieving a score of 5 in 24 of 25 cases) and melanocytic nevi (achieving a score of 4 in all four cases reported as “nevus cell nevus”). There was some variation in squamous cell carcinoma and actinic keratosis cases, with scores of 3 and 4 in 15 of 21 cases. However, non-tumor/inflammatory conditions and re-excision cases without residual tumor showed low consistency and accuracy scores, with 15 out of 25 cases scoring between 0 and 1. Notably, the two melanoma cases received a score of 0. We suspect that the poor performance in melanoma cases is due to the fact that HistoGPT was not trained on enough melanoma examples of different stages (n = 167, see Fig. 3A).

The independent clinical evaluations confirm our previous findings that HistoGPT performs well in common diseases and worse in rare diseases (see Fig. 3A, 3E, and Fig. 4D, 4E). Thus, as with all machine learning algorithms, the quality of the output is limited by the quality of the training data rather than the model architecture. In other words, HistoGPT will most likely benefit from training on larger and more diverse datasets.

## Discussion

With HistoGPT, we present a vision language model that generates pathology reports from multiple full-resolution, gigapixel whole slide images. The generated reports are of high quality for common diseases, consistent with both ground truth and independent expert review. HistoGPT surpasses the state-of-the-art foundation model GPT-4V, which is considered a powerful tool in many medical applications^18,40,41^. In addition, HistoGPT robustly predicts disease subtypes, as validated in geographically diverse cohorts. Using different prompts (e.g., “The tumor thickness is”), the model output can be tailored to specific needs. This zero-shot capability rivals existing zero-shot learning approaches based on CLIP and SigLIP. Advanced methods such as Ensemble Refinement allow us to explore the probability space of possible medical outcomes. In particular, HistoGPT’s output text is fully interpretable using attention maps that project each word in the generated report to the corresponding regions in the image.

HistoGPT was trained on 6,705 clinical cases, which is about the number of cases a pathologist in Germany must have seen to qualify for the dermatopathology examination^42^. However, this number is small by LLM standards, where models are typically trained on billions of image-text pairs from the Internet. This means that HistoGPT has probably not seen enough training signals to generate detailed reports for all scenarios. Thus, it performs worse on inflammatory diseases, which make up all minority classes, than on common classes like basal cell carcinoma, where even subtyping works in a zero-shot fashion.

Nevertheless, our work provides evidence that vision language models can be trained with significantly less data and still perform well. Although current neural networks can already predict tumor thickness or tumor subtypes with good accuracy, as has been shown in particular for basal cell carcinoma^43–49^, they require a large amount of high-quality, precisely annotated data for training and are not flexible enough to be used for tasks other than those for which they were trained. That is, these models are fully supervised and do not operate in a zero-shot fashion. Specifically for tumor thickness prediction, the above approaches are not end-to-end deep learning systems. Users must first train a segmentation model to segment the tumor region, and then use a hand-crafted mathematical algorithm to calculate tumor thickness^43–49^. HistoGPT does not require this multi-step approach because it has already learned to understand the concept of tumor thickness and more by looking at a sufficient number of image-text pairs.

So far, our model has only been trained and tested on dermatological samples. Thus, it cannot yet be generalized to pan-cancer diagnosis. In addition, our training dataset suffers from severe class imbalance, which limits its usefulness for minority classes. This problem can be partially mitigated by either “Expert Guidance” or “Classifier Guidance”. However, guidance also has its limitations, as the generated reports tend to be of higher quality when the model’s diagnostic prediction is also correct (see Supplementary Figure 1). While we have evaluated HistoGPT in real-world medical cohorts and clinical evaluations, only a large-scale clinical trial can confidently quantify the impact of HistoGPT on patients. This is beyond the scope of our current work as we are pioneering the first tool in this direction.

An interesting research direction is to fine-tune HistoGPT as a conversational chatbot using Reinforcement Learning with Human Feedback (RLHF). This will be challenging in practice, as there are currently no slide-level question-answer pairs for the model to learn from. A clinically relevant question is whether a tumor has been removed as a whole or whether there is still a tumor mass at the margins, a task that should be addressed in follow-up studies. Another clinically important question is whether and how to differentiate between primary tumors and metastases. Admittedly, indications such as tumor cell growth emanating from the epidermis can be detected with current AI. However, there will still be cases where the AI – just like human pathologists – will have difficulty making the final decision. Incorporating multimodal data, such as clinical, radiological, and sequencing data, which are often used as complementary tests in cases where pathology alone is inconclusive, is a key challenge for follow-up models.

## Methods

### Datasets

#### Munich cohort

All 15,129 histology specimens from the Munich cohort were processed and stained (with hematoxylin and eosin) at the Department of Dermatology, Technical University of Munich. They were scanned with a 20x objective at 0.173 micrometers per pixel at the Core Facility Imaging at Helmholtz Munich. All slides were fully anonymized. 100 random cases are provided in the Supplementary Material along with the reports. An example report (translated from German to English using a machine translation model) reads: “Final diagnosis: Scar. Microscopic findings: A wedge-shaped excidate with compact massive orthohyperkeratosis, focally regular acanthosis of the epidermis with hypergranulose, focally clearly flattened epidermis with elapsed reticles is presented. Underneath densely packed, partly hypereosinophilic cell-poor collagen fiber bundles, vertically placed capillary vessels. In the depth more homogenised hypereosinophilic proliferating collagen fiber bundles. Critical findings: Hypertrophic, keloid-like scar. Partial excision.”

#### Münster cohort

All 1,300 histologic samples of the Münster cohort were processed and stained (with hematoxylin and eosin) at the Department of Dermatology, University Hospital Münster. They were scanned with a 20x objective at 0.46 micrometers per pixel using a Hamamatsu NanoZoomer S360 MD at the Department of Dermatology, University Hospital Münster. The cohort includes 300 cases of three BCC subtypes (superficial, solid/nodular, infiltrating) with 100 samples each and 1000 cases from daily routine without special selection. All slides were fully anonymized. An example report (AI-translated from German to English) reads: “Lichen planus-like keratosis (regressive solar lentigo/flat seborrheic keratosis), no evidence of basal cell carcinoma in the present biopsy.”

#### Mayo cohort

A random subset of skin biopsies from a one-week period at a tertiary medical center dermatology clinic that is part of the Mayo Clinic system was blindly selected for this study. The selection included 52 retrospective dermatopathology cases consisting of 84 specimens. These cases were previously diagnosed in a digital pathology environment by board-certified dermatopathologists with more than ten years of independent practice in academic centers. Slides were scanned using a standard whole slide scanner, and whole slide images were viewed on a digital pathology image viewing platform in widespread use at the contributing authors’ institution. An example report reads: “Skin, right melolabial fold, punch biopsy: Infiltrating basal cell carcinoma with variably clear cell features, lateral biopsy edge involved, see comment COMMENT: The carcinoma is confirmed by positivity to CK903.”

### Image preprocessing

We treat all whole slide images (WSIs) belonging to a patient as one input. In other words, we have patient-level samples instead of slide-level or even patch-level data points. For CTransPath, the WSIs were downsampled 4 times, tessellated into non-overlapping patches of 256 x 256 pixels, and resized to 224 x 224 pixels using the Python library SlideIO. UNI was trained at higher resolutions and larger patch sizes. Thus, we downsampled the WSIs 3 times and used image patches of 512 x 512 pixels. Background images were detected and excluded using RGB thresholding and Canny edge detection. The inputs were then converted to PyTorch tensor objects and normalized with a mean of (0.485, 0.456, 0.406) and a standard deviation of (0.229, 0.224, 0.225). We used this specific image size and normalization parameter according to the configurations of these pre-trained vision models.

### Model architectures

We use CTransPath^22^ as our pre-trained vision encoder for HistoGPT-S and HistoGPT-M to extract 768-dimensional feature vectors for each image patch and concatenate them along the sequence dimension to obtain a matrix of size n x 768, where n is the number of image patches. The inputs are then fed into the Perceiver Resampler^32^, which is borrowed from the vision language model Flamingo^33^ with randomly initialized weights. We change the default number of latents from 64 to 640 because WSIs are much larger than natural images and require a larger dimensional latent space to store the additional information. We keep the output size of 1536 because this has been shown to work well^33^. The fixed-size outputs of dimension 640 x 1536 are then used as keys and values in the tanh gated cross-attention block (XATTN). The query vectors come from the pre-trained language model BioGPT^20^. In particular, we use one XATTN block after each language layer according to the high-performance configuration of Flamingo. The output layer of HistoGPT is a linear classifier over the vocabulary. For HistoGPT-L, we use UNI^23^ as our pre-trained vision encoder, which returns feature vectors of dimension 1024 instead of 768. We use a graph convolutional network (GCN)^34^ to encode the relative position information of each patch (given by the local neighborhood) and add it to the original feature vectors. In addition, we skip the Perceiver Resampler and replace it with the identity function. This way, we do not lose any information due to compression, as is the case with resampling.

We compare HistoGPT with HistoCLIP and HistoSigLIP. They use the feature mean of the pre-trained Perceiver Resampler as the image representation and the EOS token of the pre-trained BioGPT as the text representation. A contrastive loss then aligns both feature vectors in the common embedding space. For HistoCLIP we use the same loss as for CLIP^50^. For HistoSigLIP we use the loss proposed in SigLIP^51^. To improve performance and avoid training instabilities, we freeze the vision encoder during training. This technique is called locked-image text tuning^52^). We also compare HistoGPT with the patch-based foundation model PLIP using the contrastive pre-trained model via the provided API. To aggregate the patch-level results to the slide-level, we evaluate PLIP^16^ using the majority voting system of the related model MI-Zero^15^.

Since BioGPT and many other popular LLMs are all pre-trained on mostly English text, we need to translate the German reports into English to take advantage of their capabilities. For the translator, we choose a standard machine translation model based on the Transformer encoder-decoder architecture^30^ with the checkpoint “Helsinki-NLP/opus-mt-de-en” available on Hugging Face.

### Model training

We pre-train the Perceiver Resampler in a fully supervised manner by predicting the final diagnosis using a linear classifier on top of the slide encoder. Since the labels are provided at the patient level, this approach is also known as multiple instance learning (MIL). The classification head is then discarded and the resampler is plugged into the vision language model. We freeze all layers of HistoGPT except the cross-attention blocks. Our generative training is based on causal language modeling: Given an input, we mask the next tokens and let the model predict them. This is done in parallel over all input tokens using an upper triangular causal attention mask. Since HistoGPT-L does not need the Perceiver Resampler, we skip the pre-training step. Instead, we apply causal language modeling directly to HistoGPT-L, first to predict the diagnosis as a text string, and then to predict the full report.

For training, we use the AdamW optimizer with betas of (0.9, 0.95), a weight decay of 0.1, and an epsilon of 1e-8. The learning rate starts at zero and warms up linearly over 10 epochs to 1e-4 before decaying tenfold according to a cosine annealing scheduler. We use a gradient accumulation of 32 to simulate a larger batch size. Each training stage consists of 100 epochs using mixed precision training and gradient clipping to a Euclidean norm of 1.0. For contrastive learning, we use standard hyperparameters^50,51^.

During training, we randomly augment the text inputs to avoid overfitting common words and phrases. This is done beforehand using GPT-4 to sample 9 paraphrased texts with a temperature of 1.0 and nucleus sampling of 1.0. The prompt used is: “Rewrite the following text but be as accurate and faithful as possible to the original. Do not add or remove any information! Also, do not change the phrases ‘Microscopic findings:’ and ‘Critical findings:’, but leave them as they are.”

### Classifier guidance

We enable class imbalance awareness in HistoGPT by using a lightweight and specialized classification model. The classifier predicts one-hot encoded class indices, which are converted to text strings using a lookup table and inserted into HistoGPT. Suppose the training set contains C classes. Assume that at inference time we face a classification problem with c classes, where c ⊂ C. We extract features from each training sample with a pre-trained Perceiver Resampler and fit a classifier (either a linear layer or a full-sized model) that predicts these c classes. With this approach, we reduce the 167-class classification problem to a more tractable subset of classes. For BCC vs. ¬BCC, we consider all samples that are not BCC to be ¬BCC and fit an MLP with 100 neurons. For Melanoma vs. ¬Melanoma, we follow the same procedure. For all other classification tasks, we only train on the specific subset. For example, if we want to classify BCC vs. SCC vs. AK vs. SK, we train a classifier only on the BCC, SCC, AK, and SK training features and ignore the remaining classes. Some datasets (Queensland and Linköping) provide only annotation masks as labels. They may contain different disease labels for different regions in the same slide. In this case, we consider the prediction of one of the ground truth classes as accurate.

Simple “Classifier Guidance” does not work for HistoGPT-L because it does not use a slider encoder to which we can apply a linear classifier. However, the model can be prompted with another separate/dedicated classification model, such as TransMIL or TransformerMIL. Alternatively, we can restrict the output dictionary of the language head to the classes we are interested in and get a similar performance as with “Classifier Guidance”).

### Interpretability maps

For the HistoGPT saliency maps, we use partial derivatives and associate the output latents of the Perceiver Resampler with the corresponding input vectors. We then weight the image features with the text features using the cross-attention scores. This gives us a gradient attention map. It shows which word in the generated report corresponds to which region in a WSI. For example, we can highlight where the model sees basal cell carcinoma, how it detects tumor-infiltrating lymphocytes, and which regions it considers when measuring tumor thickness. In this way, we provide a novel approach to explainable AI by aligning visual and linguistic information.

The output of the Perceiver Resampler consists of 640 latent vectors. We compute the gradients of these latents with respect to the input patches with backpropagation. Thus, the gradient G has the form num_patches x num_latents. It tells us which image tokens have the most influence on which latent feature. The mean along the latent sequence thus gives us the most important image regions according to the vision resampler. How can we use this information to determine which of these regions corresponds to which word? One idea is to give higher weights to the latents corresponding to the words we are interested in. We get these weights by looking at the cross-attention scores of the last XATTN layer. The attention matrix A has a dimension of num_tokens x num_latents. Thus, given a target word, we can identify the corresponding target tokens and use the corresponding rows in the attention matrix as weights. Overall, the proposed *Gradient x Attention* map is given by the weighted mean

(G^T^ ∘ A[target_tokens, :].mean(dim=0)^T^)^T^.mean(dim=1).

### Evaluation metrics

We introduce two other non-trivial baselines: given the ground truth, compare two random reports with two arbitrary diagnoses (lower baseline), and compare two random reports with the same diagnosis (upper baseline). The logic behind this approach is straightforward. Medical texts often follow a structured format with a similar writing style, typically including a general description of the specimen and frequent use of common technical terms. In addition, certain diseases manifest homogeneously across patients, resulting in nearly identical report descriptions within a patient group. In such cases, the few unique terms in the reports become critical in distinguishing between different diagnoses. Therefore, these two baseline comparisons provide reference points for measuring the overall performance of our models.

Evaluating the reports generated by HistoGPT is a non-trivial task. Popular evaluation methods for natural language generation such as BLEU-4^53^, ROUGE-L^54^, and METEOR^55^ primarily compare n-grams between two documents and may not effectively capture semantic similarities. In fact, two texts may describe the same phenomena in two different ways, making a word-by-word comparison unfair. Therefore, we focus on two different quantitative performance measures: keyword overlap and sentence similarity. For the former, we use a comprehensive glossary of human-curated dermatological vocabularies^56^ to extract important medical keywords from the ground truth notes. In addition, we use ScispaCy^36^, a biomedical named entity recognition (NER) tool, to capture a broader range of technical terms. We then determine how many keywords from the ground truth text can be found in the generated text. The Jaccard index is used to quantify their overlap. To find a match in the generated report, we use an advanced version of Gestalt pattern matching (Ratcliff and Obershelp, 1988) available in the Python library difflib. We use the default cutoff threshold of 0.6. This value strikes a balance between matching every word as a target and matching only exact overlaps. The latter is undesirable because it ignores different grammatical forms of a word. As a result, some unrelated words will inevitably be matched. In this case, the Jaccard index can be considered a relative measure, since the same approach is applied to each model.

The above measures still miss some semantic nuances because certain concepts or observations (e.g., disease characteristics, tissue subtypes, cellular characteristics) may be expressed in complex phrases, possibly even involving negations. To remedy this, we use BioBERT^37^ fine-tuned^57^ for natural language inference (NLI) and semantic textual similarity (STS) assessments. This embedding model provides the feature vectors of the generated report and the ground truth, allowing us to compute their cosine similarity as a measure of semantic understanding. To go beyond the domain-specific use of language, we apply a general large-scale embedding model, GPT-3-ADA^31^, to capture a broader range of linguistic information. Similarly, we use BERTScore^58^ to compute the syntactic relationship between generated and ground truth reports at the subword level.

For Ensemble Refinement, we summarize the bootstrapped reports by prompting GPT-4-Turbo with the instruction “Summarize the following text:”. Since sampling for Ensemble Refinement is massively time-consuming and relies on an expensive API call, we only compute the scores on a random subset of the test set (10%). However, the standard deviation among the samples remains similar to the models on the full test set, indicating that the final score would not change much.

## Data availability

The datasets are either publicly available at the link provided or can be requested from the original investigators:

- CPTAC (https://www.cancerimagingarchive.net/collection/cptac-cm/)
- Linköping (https://datahub.aida.scilifelab.se/10.23698/aida/drsk)
- Queensland (https://espace.library.uq.edu.au/view/UQ:8be4bd0)
- TCGA (https://portal.gdc.cancer.gov/projects/TCGA-SKCM)

## Code availability

The code for the model can be found at https://github.com/marrlab/HistoGPT. The weights for the model are available at https://huggingface.co/marr-peng-lab/histogpt.

## Acknowledgments

M.T., S.W.J., and V.K. are supported by the Helmholtz Association under the joint research school “Munich School for Data Science – MUDS”. C.M. acknowledges funding from the European Research Council (ERC) under the European Union’s Horizon 2020 research and innovation program (Grant Agreement No. 866411) and support from the Hightech Agenda Bayern.

We thank the dermatopathologists R. Hein, R. Franz, S. Möckel, A. Steimle-Grauer, C. Andres, and S. Roenneberg (Munich) for the detailed review of the patient material and for providing the data for this project. We also thank Nina Witte (Münster) for her technical and organizational support.

## Author information

M.T. developed the methods, implemented the code, and performed the experiments. P.S. provided domain knowledge, collected the dataset, and evaluated the quality of the reports. S.J.W. helped with the experiments by providing the MIL and zero-shot learning results. V.K. managed the data processing pipeline through patching and feature extraction. B.N. was responsible for the experiments at the Mayo Clinic. V.L. curated the data and trained the MIL models. A.F. processed and scanned the whole slide images from the Munich cohort. A.B. collected and annotated the internal training dataset with images and reports. R.K. provided medical advice and designed the real-world evaluation metrics. T.B. provided resources for data acquisition and contributed to the drafting of the manuscript. N.I.C. helped in the pathology analysis at the Mayo Clinic. R.G. performed the pathology analysis at the Mayo Clinic. W.C. supervised the project for the Mayo Clinic cohort. K.E. supervised the study and the compilation of the internal Munich dataset. S.A.B. supervised the compilation of the external Münster data set and assessed the quality of the report. T.P. supervised the machine learning approach. C.M. supervised the study.

## Ethics declaration

### Ethics statement

All research procedures were conducted in accordance with the Declaration of Helsinki. Ethical approval was granted by the Ethics Committee of the Technical University of Munich (reference number 2024-98-S-CB) and the Ethics Committee of Westfalen-Lippe (reference number 2024-157-b-S).

### Inclusion and diversity

An interdisciplinary team of computer scientists, dermatologists, and pathologists from several institutions worked closely together. They shared their expertise and maintained the integrity of the scientific record throughout the study. Local researchers were involved in the research process to ensure that the study was locally relevant. International researchers were also involved to ensure that the study was intentionally relevant. Roles and responsibilities were agreed upon before the study and resources were allocated accordingly.

## Supplementary Figures

**Supplementary Figure 1.**
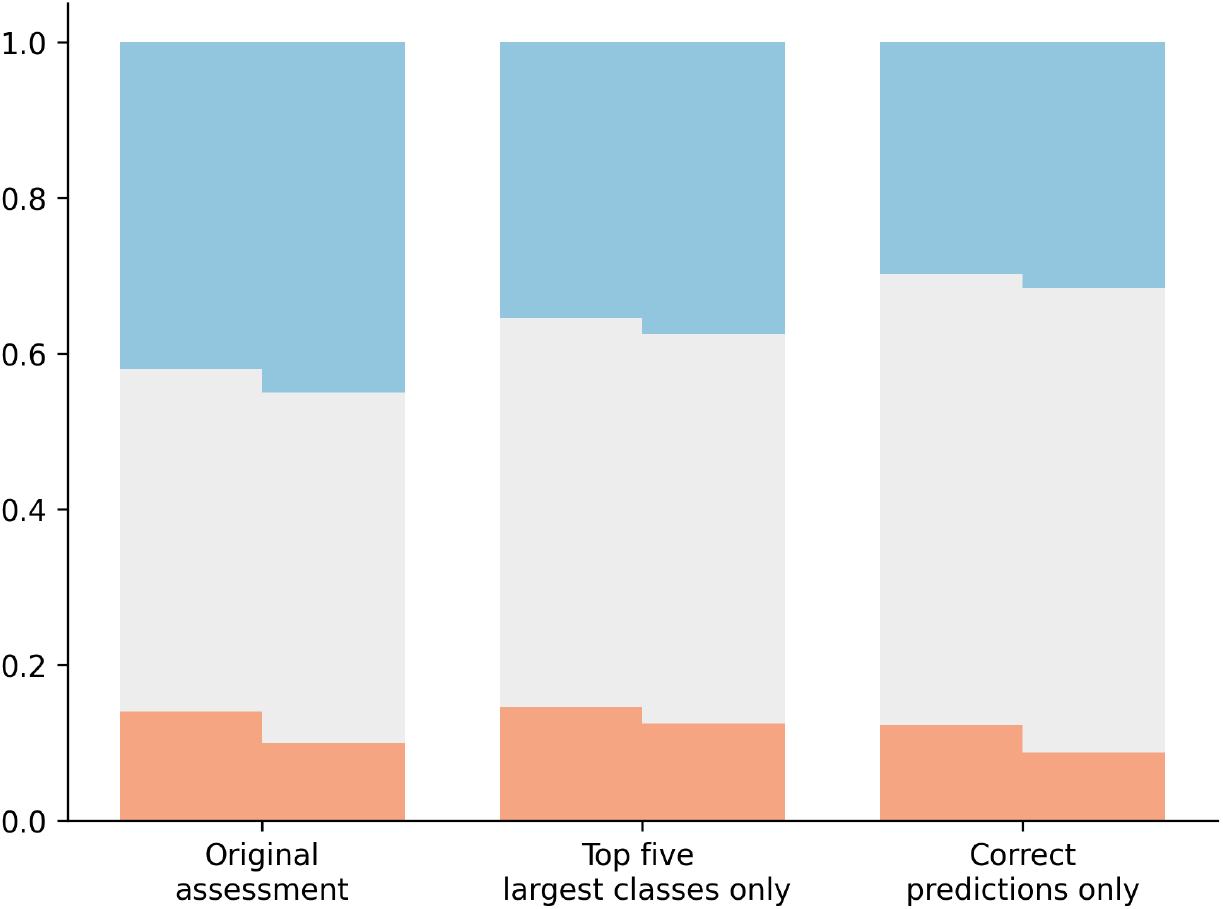
Results for the blinded study where two pathologists evaluate the performance of the AI-generated reports against the human reports. We filter the results for the five largest classes and for cases where the model’s prediction matches the ground truth.

**Supplementary Figure 2.**
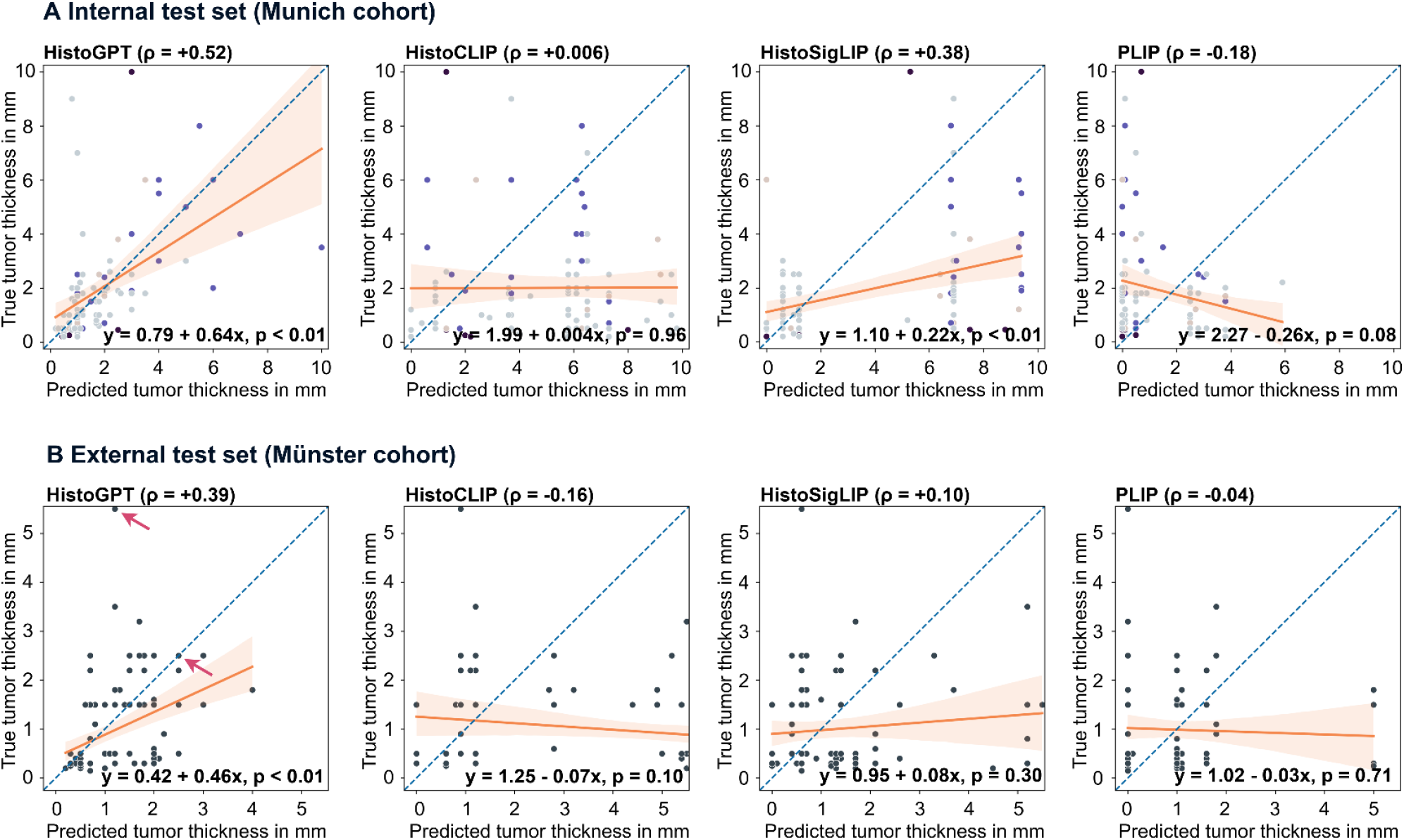
Zero-shot prediction of tumor thickness by HistoGPT-M, HistoCLIP, HistoSigLIP, and PLIP. (A) Results are summarized in a scatter plot with a regression line for the Munich cohort. All data points are color-coded according to Figure 3A. (B) A similar plot is generated for the Münster cohort.

**Supplementary Figure 3.**
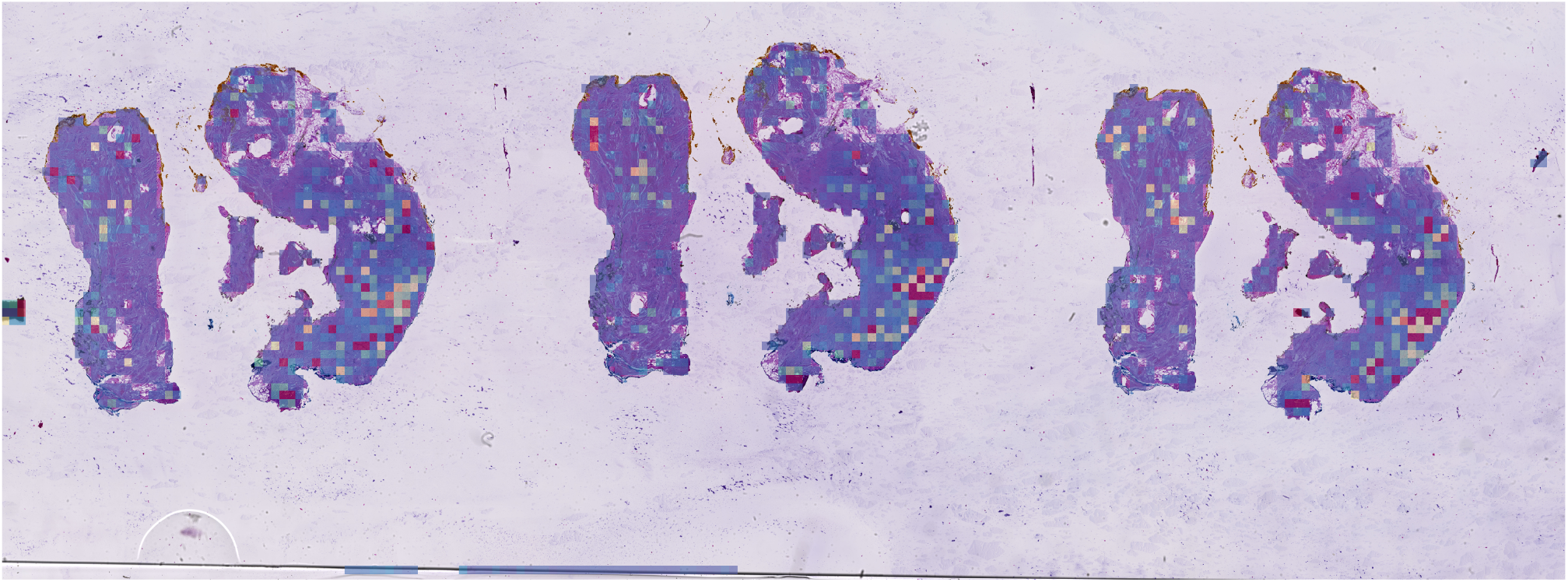
This basal cell carcinoma specimen is missing large portions of the epidermis. Thus, the model did not find a reference point to orient the slide, which likely caused the overestimation of thickness. The ground truth measurement is 1.8 mm and the model prediction is 4.8 mm.

**Supplementary Figure 4.**
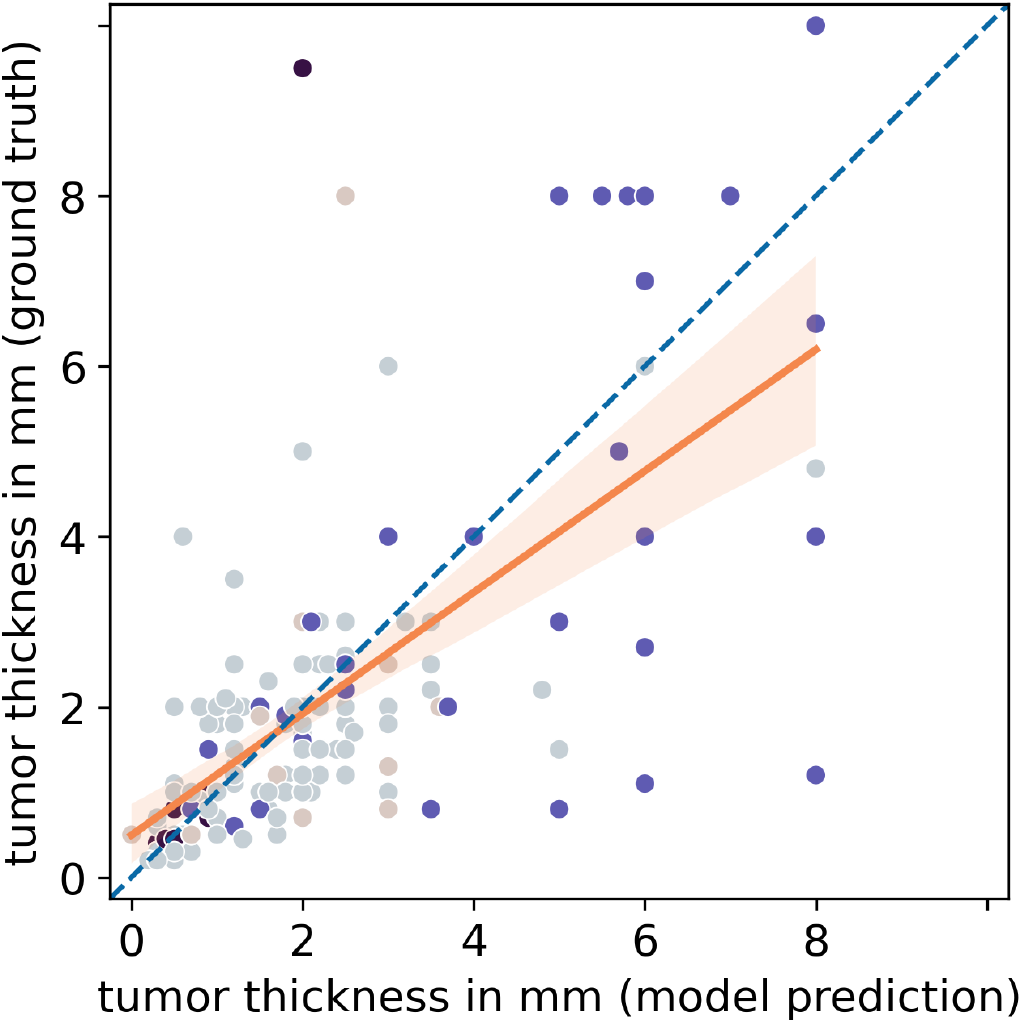
Zero-shot prediction of tumor thickness by HistoGPT-L on the internal Munich test split.

**Supplementary Figure 5.**
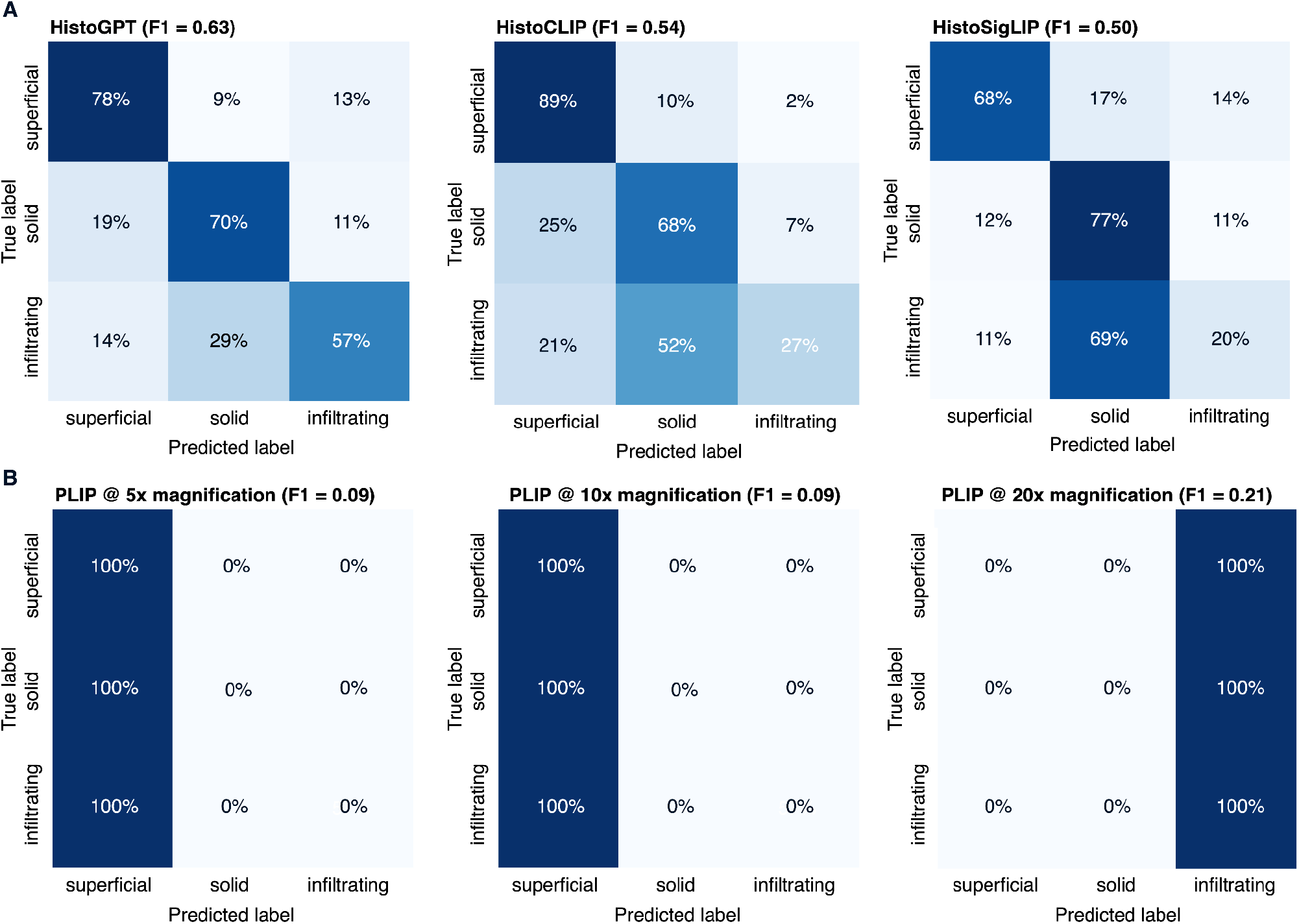
Zero-shot tumor subtype prediction results summarized as a confusion matrix for HistoGPT-M, HistoCLIP, HistoSigLIP, and PLIP.

**Supplementary Figure 6.**
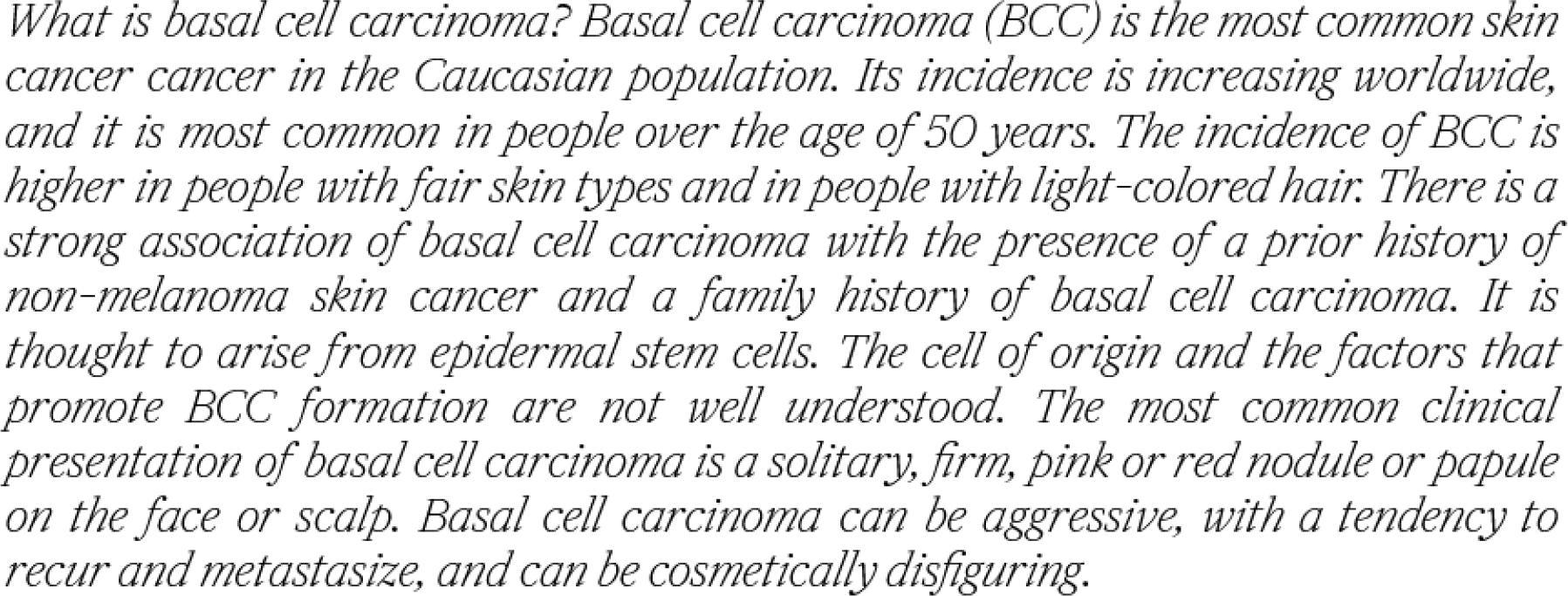
Since the BioGPT language model is frozen during training, HistoGPT can be easily converted to a language-only model by taking only text as input, while retaining all the capabilities of the pre-trained BioGPT. Above we see the Ensemble Refinement output for the definition of basal cell carcinoma (BCC).

## Supplementary Tables

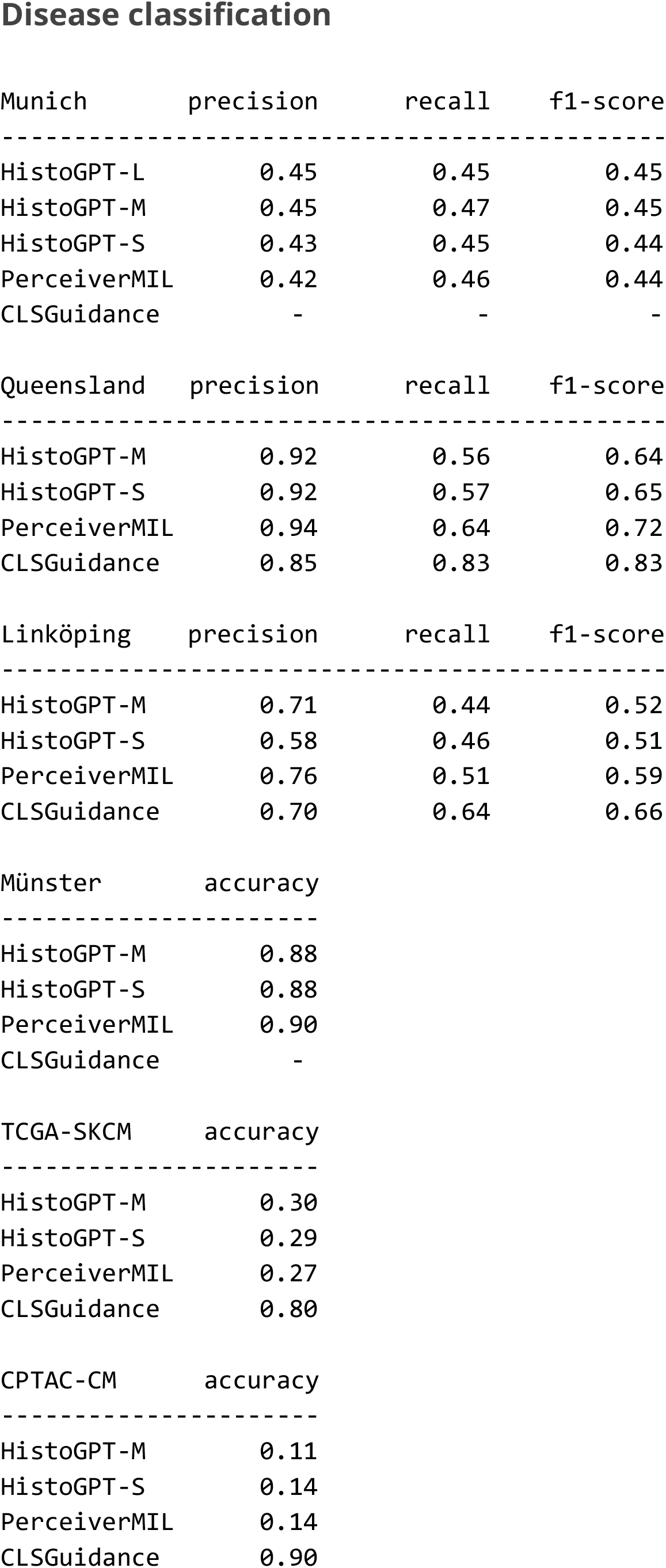
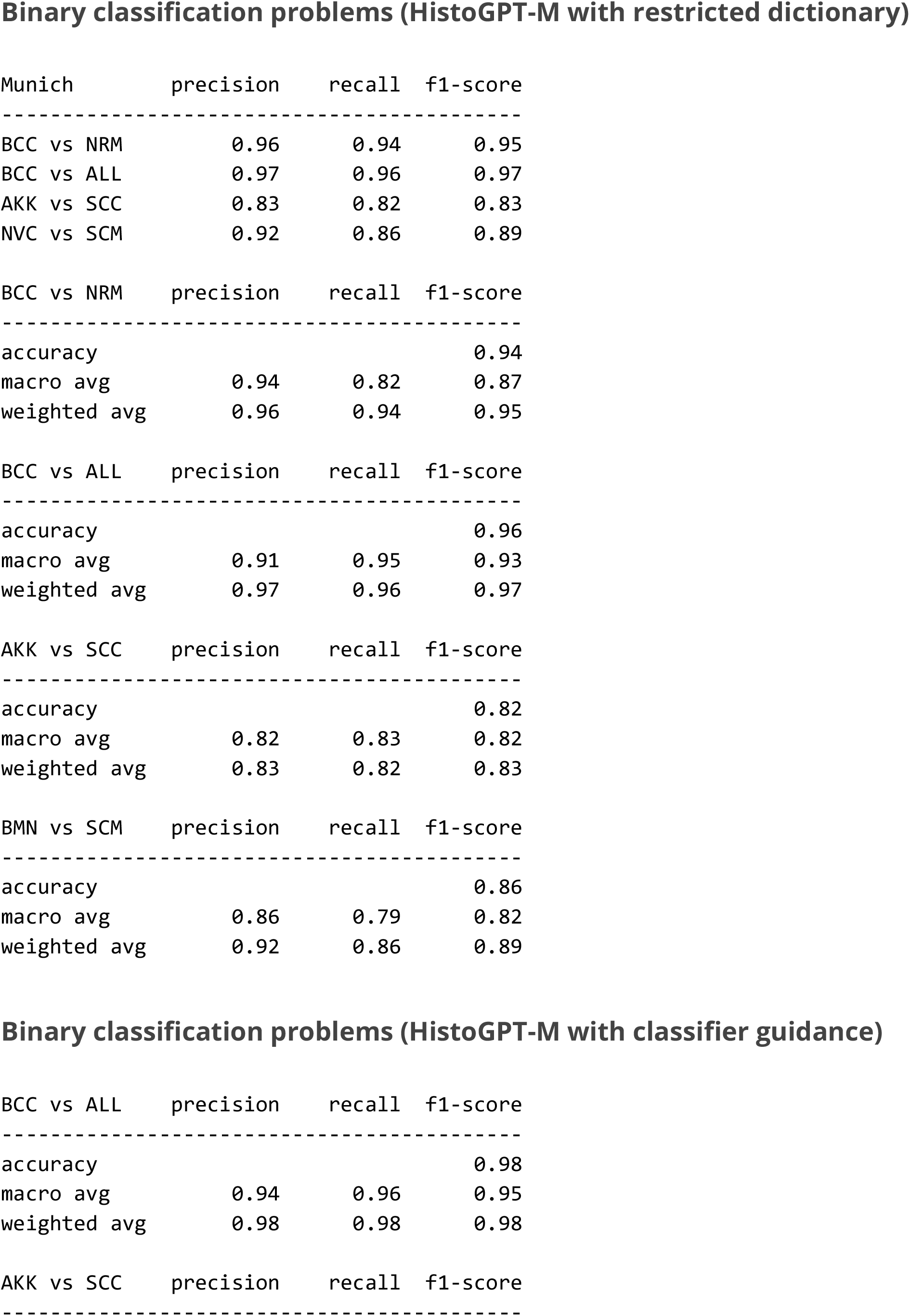
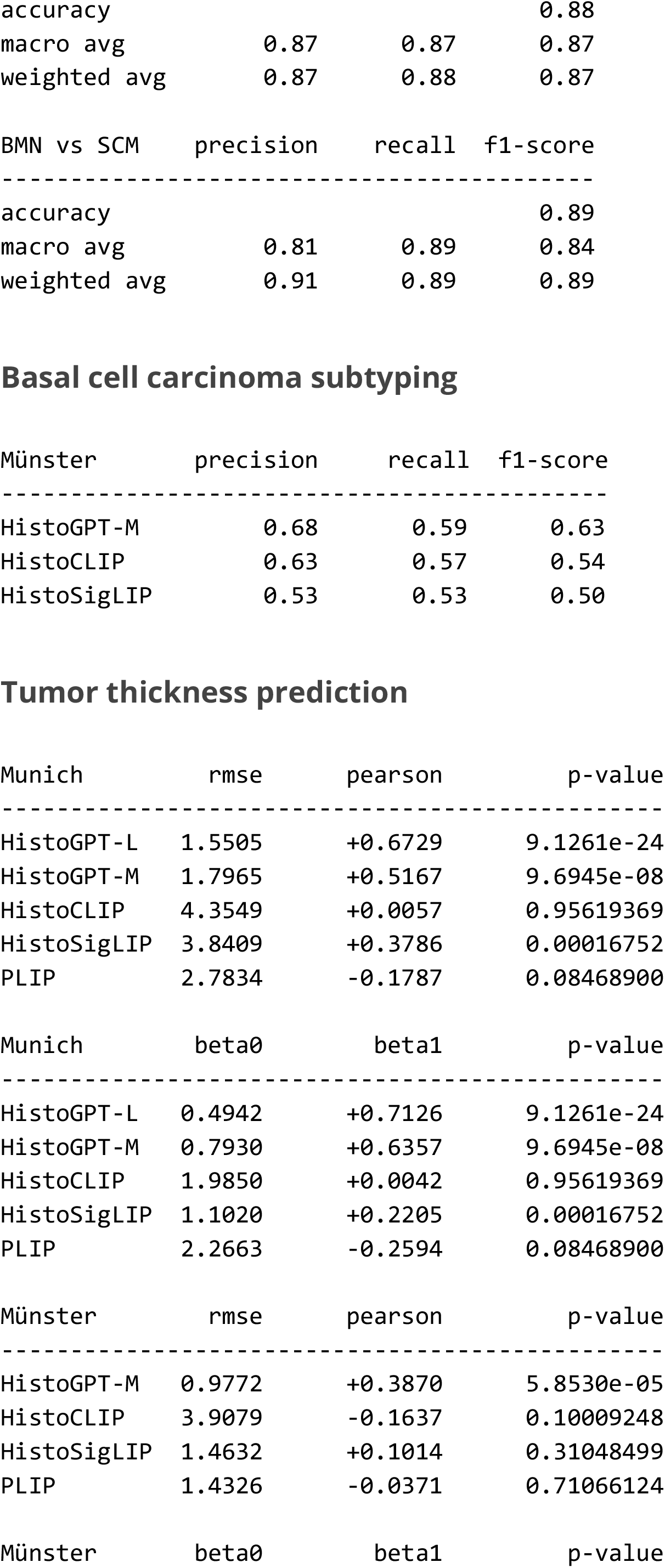
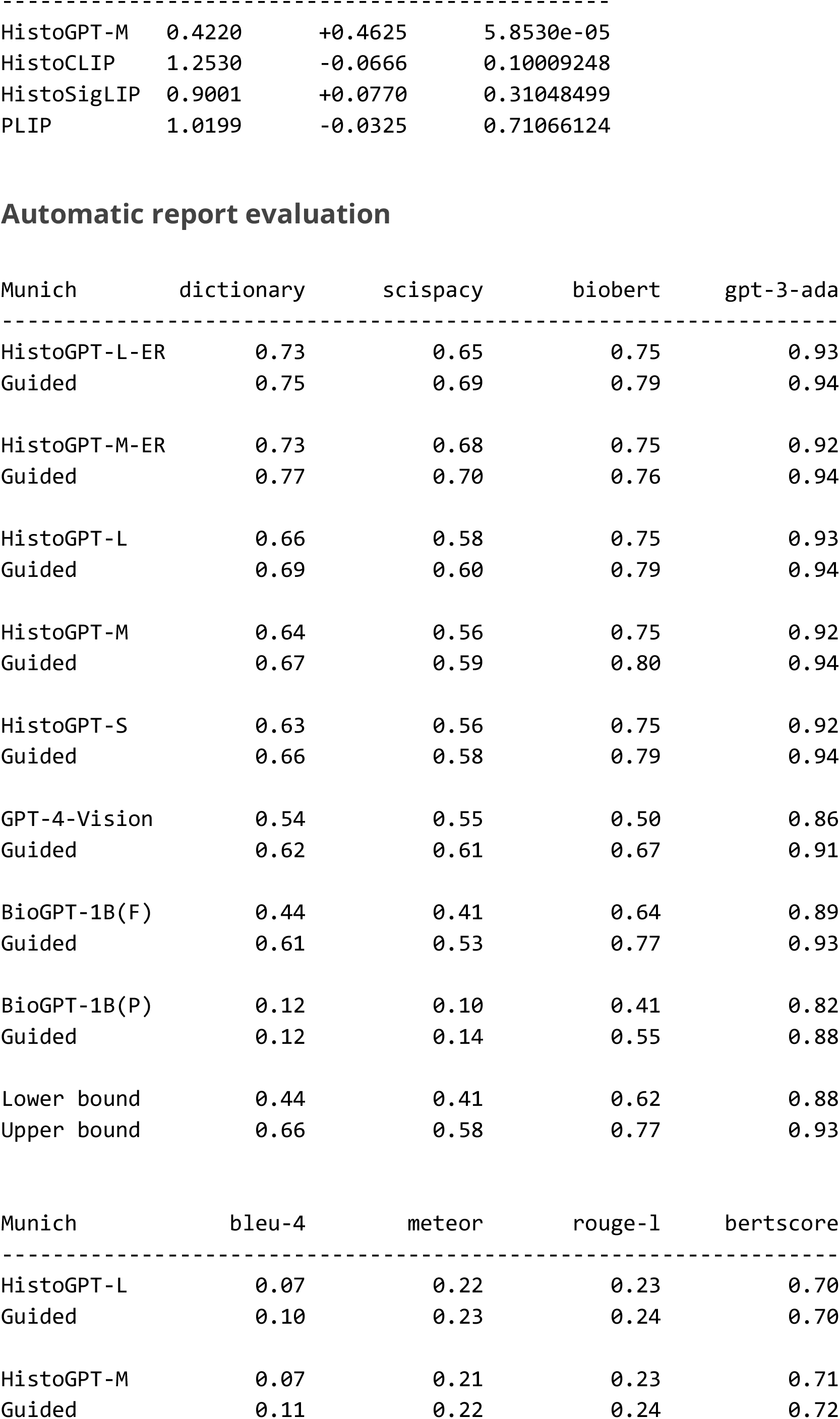
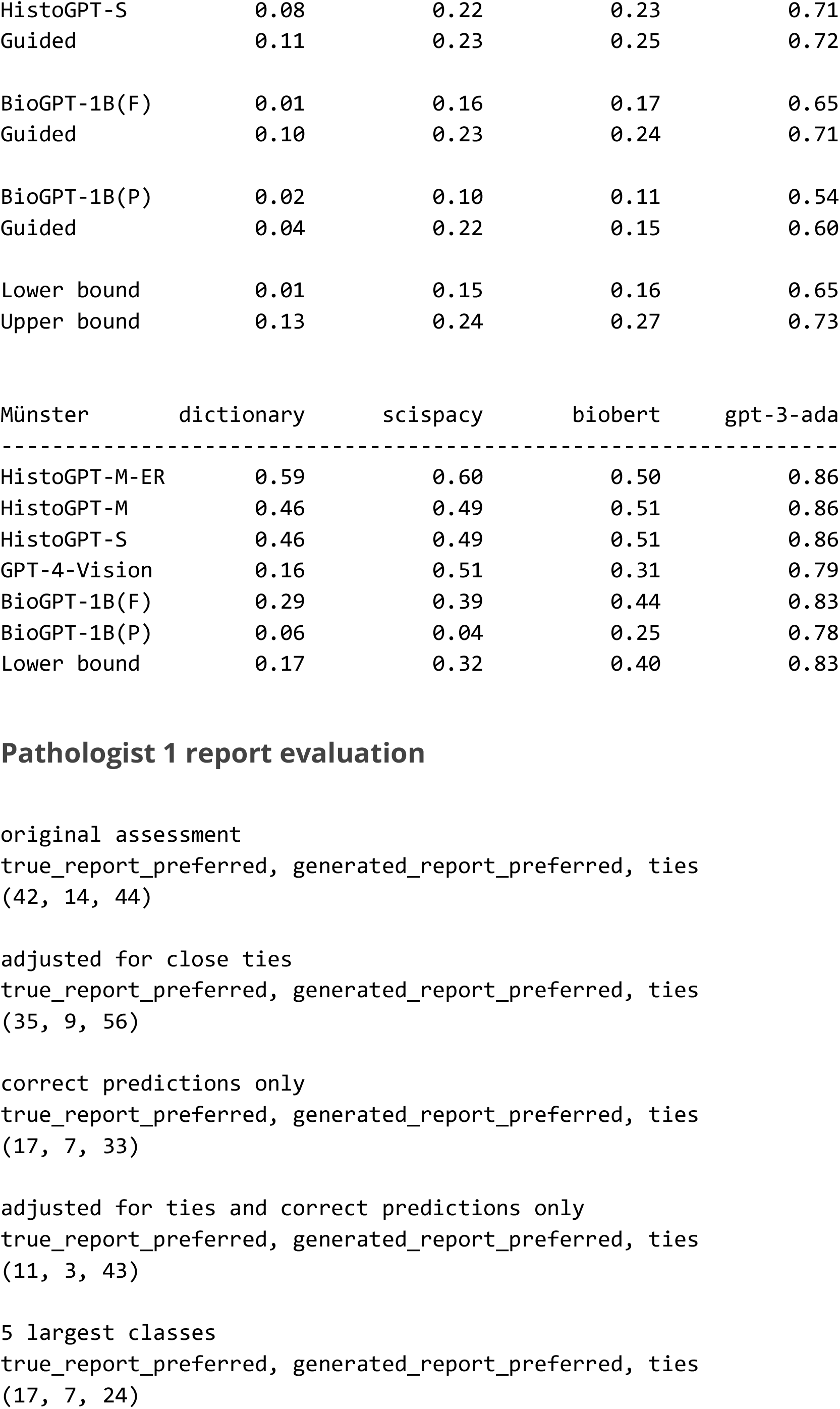
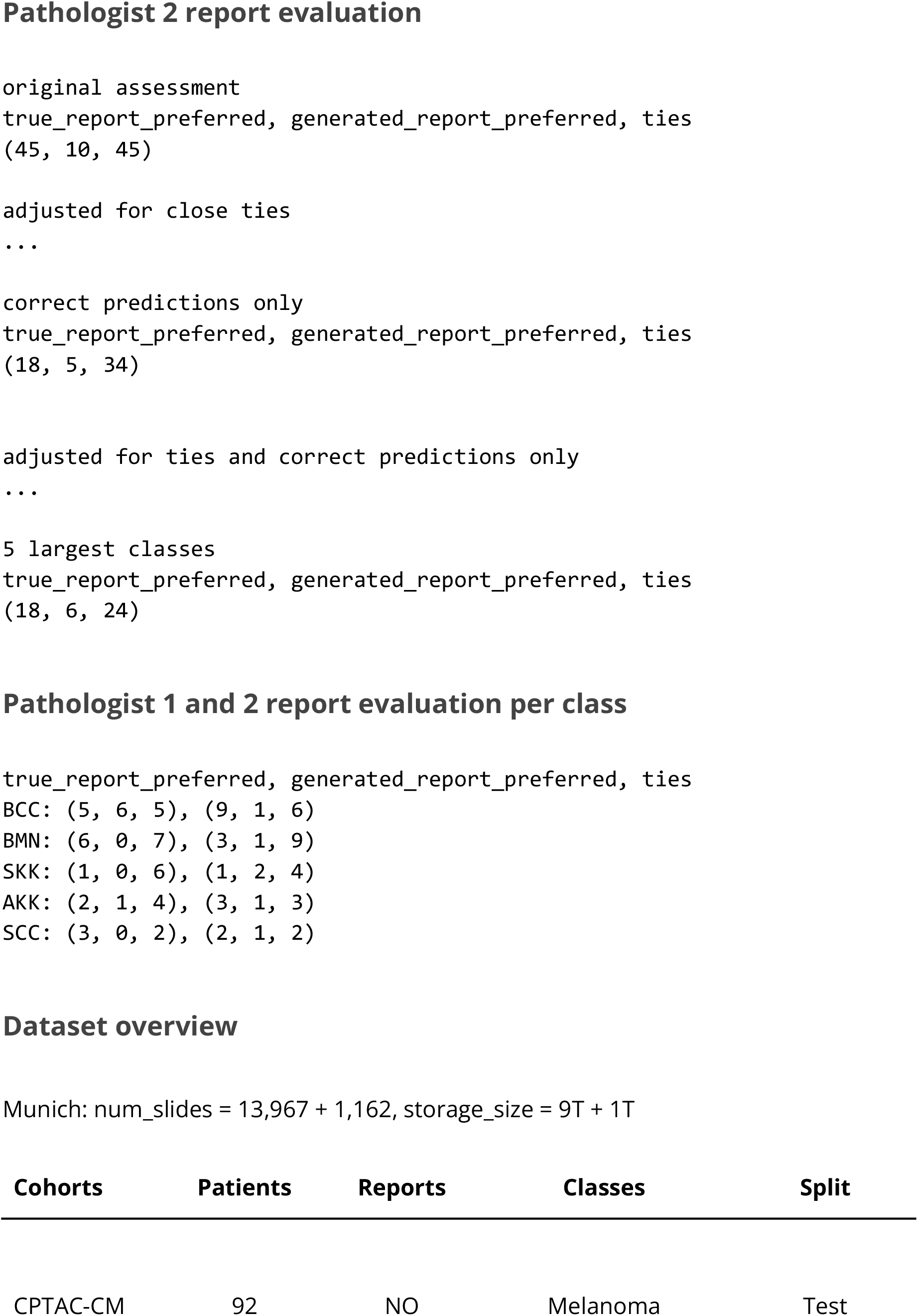
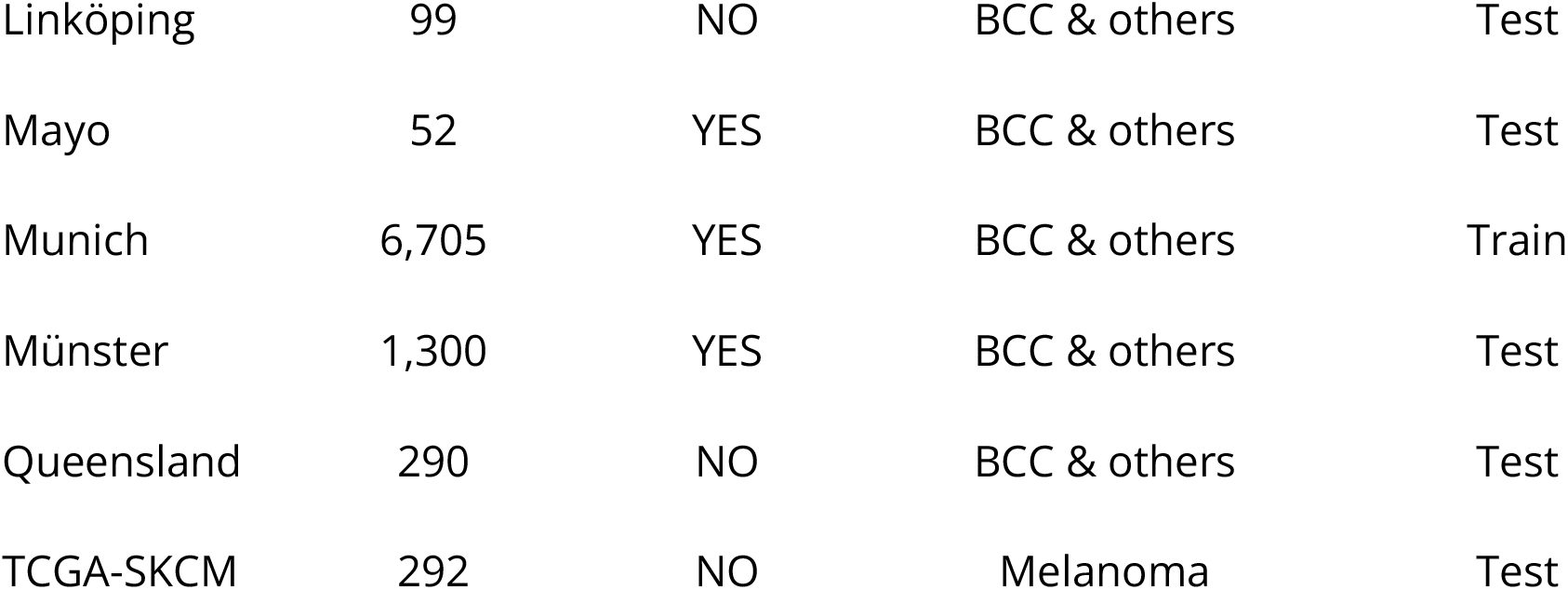

